# A mechanistic neural network model predicts both potency and toxicity of antimicrobial combination therapies

**DOI:** 10.1101/2025.03.19.25324270

**Authors:** Harkirat Singh Arora, Katherine Lev, Aaron Robida, Ramraj Velmurugan, Sriram Chandrasekaran

## Abstract

Antimicrobial resistance poses a major global threat due to the diminishing efficacy of current treatments and limited new therapies. Combination therapy with existing drugs offers a promising solution, yet current empirical methods often lead to suboptimal efficacy and inadvertent toxicity. The high cost of experimentally testing numerous combinations underscores the need for data-driven methods to streamline treatment design. We introduce CALMA, an approach that predicts the potency and toxicity of multi-drug combinations in *Escherichia coli* and *Mycobacterium tuberculosis*. CALMA identified synergistic antimicrobial combinations involving vancomycin and isoniazid that were antagonistic for toxicity, which were validated using *in vitro* cell viability assays in human cell lines and through mining of patient health records that showed reduced side effects in patients taking combinations identified by CALMA. By combining mechanistic modelling with deep learning, CALMA improves the interpretability of neural networks, identifies key pathways influencing drug interactions, and prioritizes combinations with enhanced potency and reduced toxicity.

## INTRODUCTION

Antimicrobial resistance (AMR) occurs due to prolonged exposure to antibiotics, which enables bacteria to adapt and survive through natural selection. In the absence of effective interventions, drug-resistant infections could lead to 10 million deaths annually by 2050, potentially causing economic disruptions on a scale comparable to the 2008–09 global financial crisis [1]. Combination therapy, which involves the use of two or more drugs to treat a single illness, represents a promising strategy to address antibiotic resistance [2], [3]. However, current methodologies for designing combination therapies frequently rely on empirical approaches, which may result in suboptimal efficacy and increased toxicity risks [4]. Furthermore, the combinatorial explosion drastically increases the search space for effective combinations, even for a single bacterial strain. Consequently, experimentally testing all possible combinations is a time-consuming and resource-intensive process [5], [6].

Recent advancements in artificial intelligence (AI) methods have accelerated the search for potent drug combinations. For example, the INDIGO approach employed chemogenomic profiles of individual drugs to predict the potency scores of drug combinations and accurately identify approximately 60% of the top synergistic and antagonistic pairwise drug combinations against *E. coli* [7]. The application of INDIGO was further extended to predict drug combination potency scores in *M. tuberculosis* using transcriptomics data, with predicted outcomes correlating with clinical efficacy [8]. The recently developed CARAMeL methodology accounted for the influence of metabolic environments on drug combination potency [9].

Despite their efficacy in reducing the search space for drug combinations against AMR, these methods focus only on synergy against the pathogen and do not account for potential toxic interactions among the drugs. Further, they are based on traditional machine learning (ML) algorithms and do not provide insights into pathways associated with drug response and the impact of specific pathways on model predictions.

A recent study indicates that approximately 90% of drug candidates fail in clinical trials, with nearly 30% of failures attributable to unanticipated adverse side effects [10], [11]. Addressing and mitigating side effects is crucial for the successful development and market viability of new drugs. Antimicrobials, including antibiotics, represent one of the largest classes of medications associated with impaired kidney function [12], [13] and drug-induced liver injury in nearly 46% of patients [14]. Although combination therapies can have lower individual dosages to reduce toxicity, they still pose the risk of adverse and underexplored drug interactions. In the United States, multi-drug side effects are an urgent concern, affecting approximately 15% of the population and leading to significantly higher healthcare and pharmacy costs [15], [16]. Increased side effects can also exacerbate AMR by decreasing patient adherence to the full course of antibiotic therapy.

Previous approaches to designing combination therapies have typically overlooked the toxicity and safety profiles of drug combinations. Additionally, most machine-learning models for predicting toxicity focus on single-drug treatments [17]–[20]. Although recent studies have been proposed to predict the toxic effects of drug combinations using graph-based representations to enhance model accuracy, they often do not quantify the toxicity or elucidate the pathways governing drug combination toxicity and potency [21], [22].

To address these limitations, we present an approach that integrates Artificial Neural Networks (ANNs) with Genome-scale Metabolic Models (GEMs) to predict the potency and toxicity of drug combinations. GEMs are computational models that contain the entire known set of biochemical reactions encoded in the genome of an organism, including processes that are primary and secondary antibiotic targets such as central carbon metabolism, lipid synthesis, cell wall synthesis, nucleotide metabolism, and translation [23]. Metabolism plays a key role in influencing drug toxicity, potency and synergy [24]. Mechanistically incorporating GEM data into ANNs provides a systems-level perspective of the bacterial response to combinatorial antibiotic treatment and linking these responses to drug potency and toxicity. Here we focus on the gram-negative model organism *E. coli* and the pathogen *M. tuberculosis*, which infects millions worldwide annually. Of note GEM models and extensive drug interaction data are available for these organisms to benchmark our approach with prior methods.

## RESULTS

### Integration of GEMs and ANNs for Predicting Drug Combination Potency and Toxicity

Our approach, CALMA, involves a three-step methodology: (i) simulating metabolic reaction fluxes under individual drug conditions utilizing GEMs, (ii) processing individual reaction flux data to construct joint profile features that account for both the similarity and uniqueness among drugs in a combination, and (iii) developing an ANN model to predict drug combination potency and toxicity scores using these joint profile features.

GEMs are an efficient, mathematical representation of an organism’s known metabolic potential. The metabolic network can be mathematically depicted as a stoichiometric matrix, with metabolites represented along the rows and reactions along the columns (Figure 1A). Constraints such as mass balance, thermodynamics, omics data, and media conditions are incorporated into the model to simulate reaction flux, optimizing a specific objective function such as biomass.

**Figure 1.**
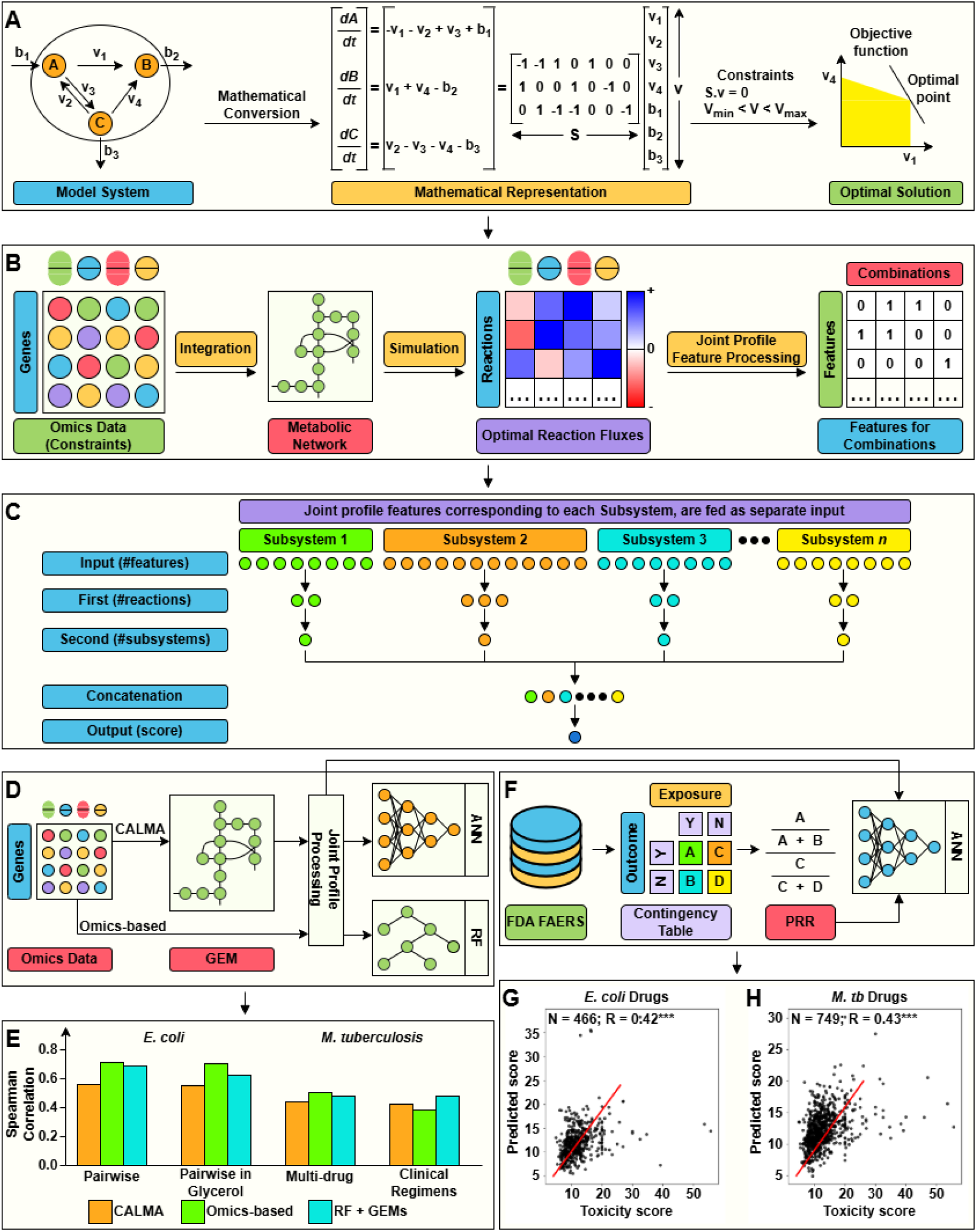
Overview and Validation of CALMA for Drug Combination Potency and Toxicity Prediction. CALMA employs an Artificial Neural Network (ANN) model integrated with data from Genome-scale Metabolic Models (GEMs) to predict drug combination potency and toxicity scores. **(A)** GEMs are computational models representing an organism’s metabolic network, detailing various reactions and metabolites, and are used to simulate reaction fluxes under different constraints and objectives. **(B)** Omics data for diverse drug and media conditions are incorporated as constraints into the metabolic network to simulate reaction fluxes, followed by joint profile processing to create combination profiles. **(C)** Features from joint profiles linked to the same subsystem are input separately into the ANN model, enabling prediction of potency and toxicity outcomes. **(D)** Schematic comparing the benchmarking of CALMA with the Omics-based approach. **(E)** Bar chart showing the Spearman rank correlation between experimental and predicted potency scores across different datasets for CALMA, Omics-based, and RF + GEMs approaches. **(F)** Overview of the development of the Drug Combination Toxicity prediction model, with GEMs and FAERS database data used to simulate reaction fluxes and provide toxicity scores (Proportional Reporting Ratio, PRR). CALMA trains the toxicity score prediction model using reaction fluxes as input. Scatterplots show drug combination toxicity prediction models **(G)** for *E. coli* and **(H)** *M. tuberculosis* GEMs, noting dataset interactions (N) and Spearman rank correlation (R).

We employed the *E. coli* GEM model iJO1366 [25] and the *M. tuberculosis* GEM model iEK1008 [26] to simulate metabolic reaction fluxes at a steady state. Chemogenomics and transcriptomics data [8] served as additional constraints for the respective *E. coli* and *M. tuberculosis* GEMs for simulating reaction flux profiles under different drug conditions. This simulation produces a reaction flux matrix, mapping reactions along the rows and drugs along the columns (Figure 1B). The individual reaction flux profiles were discretized based on differential flux activity (either positive or negative). These discretized profiles were then processed to generate joint profile features, incorporating sigma and delta scores that measure similarity of flux profiles. Through this joint profile processing, each reaction is represented by a set of four features (Methods).

The design and architecture of the ANN model is inspired by and integrated with information derived from GEMs. GEMs provide information on the mapping of each metabolic reaction to a specific subsystem. The *E. coli* GEM (iJO1366) comprises a total of 2,583 metabolic reactions mapped to 37 subsystems. Similarly, the *M. tuberculosis* GEM comprises 1,226 metabolic reactions mapped to 63 subsystems. It is noteworthy that each reaction within the GEM is mapped to a single subsystem. Ultimately, the ANN model predicts the potency and toxicity scores of drug combination therapies based on reaction fluxes from GEMs.

The ANN model comprises three components: (i) the input layer, (ii) hidden layers, and (iii) the output layer. The input layer is utilized to provide input features to the model, while the output layer generates the final output. The hidden layers perform non-linear transformations on the input features to learn patterns for output prediction. The design of these three components in CALMA is as follows:

i. Input layer: Each reaction in the joint profile features is represented by a set of four features; thus, the total number of input features is four times the number of reactions in the GEM. The number of subsystems within the GEM informs the structure of the input layer. Features corresponding to reactions within the same subsystem are grouped together as inputs to the model (Figure 1C). Therefore, the number of input sub-layers in the model equates to the number of subsystems in the GEM. For instance, the *E. coli* GEM (iJO1366) comprises 2,583 reactions mapped to 37 subsystems, resulting in an ANN model with 10,332 (4 × 2,583) features in the input layer, distributed across 37 different input sub-layers. Similarly, the ANN model for *M. tuberculosis* encompasses 4,904 (4 × 1,226 reactions) features across 63 different input sub-layers (Supplementary Tables 11 and 12).
ii. Hidden layers: Features in the input layer of the ANN model undergo further processing via non-linear transformations in the hidden layers. CALMA comprises two hidden layers in its ANN model. Input features within separate input sub-layers are processed into separate first hidden sub-layers. The number of neurons within each first hidden sub-layer is equivalent to the number of reactions within the subsystem represented by the corresponding input sub-layer. The total number of neurons in the first hidden layer equals the number of reactions in the GEM. The non-linear transformations from the first hidden layers are then processed into the corresponding second hidden layers. Each first hidden sub-layer is processed into a single neuron in the second hidden layer; thus, the total number of neurons in the second hidden layer corresponds to the number of subsystems in the GEM. The value of the neuron in the second hidden layer is a function of the input features of the corresponding input sub-layer, remaining independent of features from other input sub-layers (Figure 1C). Ultimately, the neurons in the second hidden layer are concatenated.
iii. Output layer: The neurons in the concatenated layer are processed to generate the output from the model, which can either be the potency or toxicity score of the drug combination.

### CALMA is benchmarked for predicting drug combination potency scores

To evaluate CALMA’s performance in predicting drug combination potency scores, CALMA predictions were compared against existing datasets and approaches (INDIGO and MAGENTA) [7], [8], [28] (Figure 1D). Furthermore, the performance of CALMA was also compared with a control model replacing ANNs with Random Forests (RF) (RF + GEMs).

1. We evaluated the performance of CALMA against the following datasets: 171 Pairwise drug combination outcomes among 19 drugs in *E. coli*. CALMA achieved a correlation of R = 0.56, p ≈ 10^-14 with experiments based on 5-fold CV. (Methods; Supplementary Figure 2A)
2. 55 Pairwise drug combination outcomes for *E. coli* in M9 glycerol media (R = 0.55, p ≈ 10^-6; Supplementary Figure 2B).
3. 232 Multi-way drug combination outcomes for *M. tuberculosis* (R = 0.44, p ≈ 10^-13; see Supplementary Figure 2C).
4. 57 Multi-drug TB treatment regimen efficacy in clinical trials (R = 0.42, p ≈ 10^-4; Supplementary Figure 2D).

Overall, CALMA yielded significant and accurate predictions across various datasets for both *E. coli* and *M. tuberculosis*, (Figure 1E; Supplementary Table 3). ANNs require substantial amounts of data to outperform traditional machine-learning algorithms, such as Random Forests, which can perform well with fewer data points. Although CALMA does not surpass existing methodologies that use Random Forests, the rationale for utilizing a mechanistic ANN architecture in CALMA is for enhanced interpretability without compromising performance, as evidenced by highly significant predictions comparable to existing methods, and flexibility to predict both toxicity and potency.

### CALMA predicts drug combination toxicity scores with significant accuracy

Drug discovery and development require not only the design of potent therapeutics against pathogenic cells but also treatments that are safe for human cells. We next trained the CALMA model to predict toxicity scores. Similar to the drug combination potency prediction model, the joint feature profiles derived from metabolic reaction fluxes simulated by GEMs (Figure 1B) were utilized to train the ANN model (Figure 1C) for predicting drug combination toxicity scores from the FDA Adverse Event Reporting System (FAERS). The TWOSIDES database [29], derived from FAERS, offers data in the form of drug combination-side effect pairs alongside corresponding toxicity scores, referred to as PRR (Proportional Reporting Ratio) scores. The PRR score is calculated based on the number of cases associated with the presence and absence of drug combination exposure and corresponding side effects (Figure 1F, Methods). The PRR score quantifies the occurrence of side effects in response to drug combination therapy relative to random chance. Therefore, a higher PRR score indicates greater toxicity of the combination therapy.

Since our goal is to identify pathogenic targets associated with toxicity in humans, we utilized the same set of inputs used for predicting potency; this enables direct comparison of model weights for potency and toxicity and identify selective pathway targets. We employed both *E. coli* and *M. tuberculosis* GEMs to simulate metabolic reaction fluxes for drugs for which omics data were available and which were also represented in the TWOSIDES database. This resulted in an *E. coli* dataset of 466 drug combinations and an *M. tuberculosis* dataset of 749 combinations. Following the previously described model architecture (Figure 1C), an ANN model was trained to predict drug combination toxicity scores. The model demonstrated significant performance for both datasets using ten-fold cross-validation: *E. coli* (R = 0.42, p ≈ 10^-18, Figure 1G) and *M. tuberculosis* (R = 0.43, p ≈ 10^-35, Figure 1H).

### Analysis of Drug Combination Landscapes for *E. coli* and *M. tuberculosis*

Given CALMA’s significant accuracy in predicting drug combination potency and toxicity scores, we generated a Drug Combination Landscape, a scatterplot where potency scores are represented along the x-axis and toxicity scores along the y-axis, with each point corresponding to a specific drug combination (Figure 2A). The most favourable combinations are those positioned in the bottom-left section of the landscape and correspond to combinations with low interaction scores (synergistic) and low PRR scores (less toxic). This landscape not only facilitates the assessment of a combination’s safety and effectiveness but also reduces the search space for potential drug combinations. Combinations that fall into both the top 15% categories for potency and toxicity are highlighted in blue on the drug combination landscapes (*E. coli*, Figure 2B; *M. tuberculosis*, Figure 2C). The predicted drug combination potency and toxicity scores for all 595 combinations in *E. coli* and *M. tuberculosis* are provided in Supplementary Tables 6 and 7, respectively.

**Figure 2.**
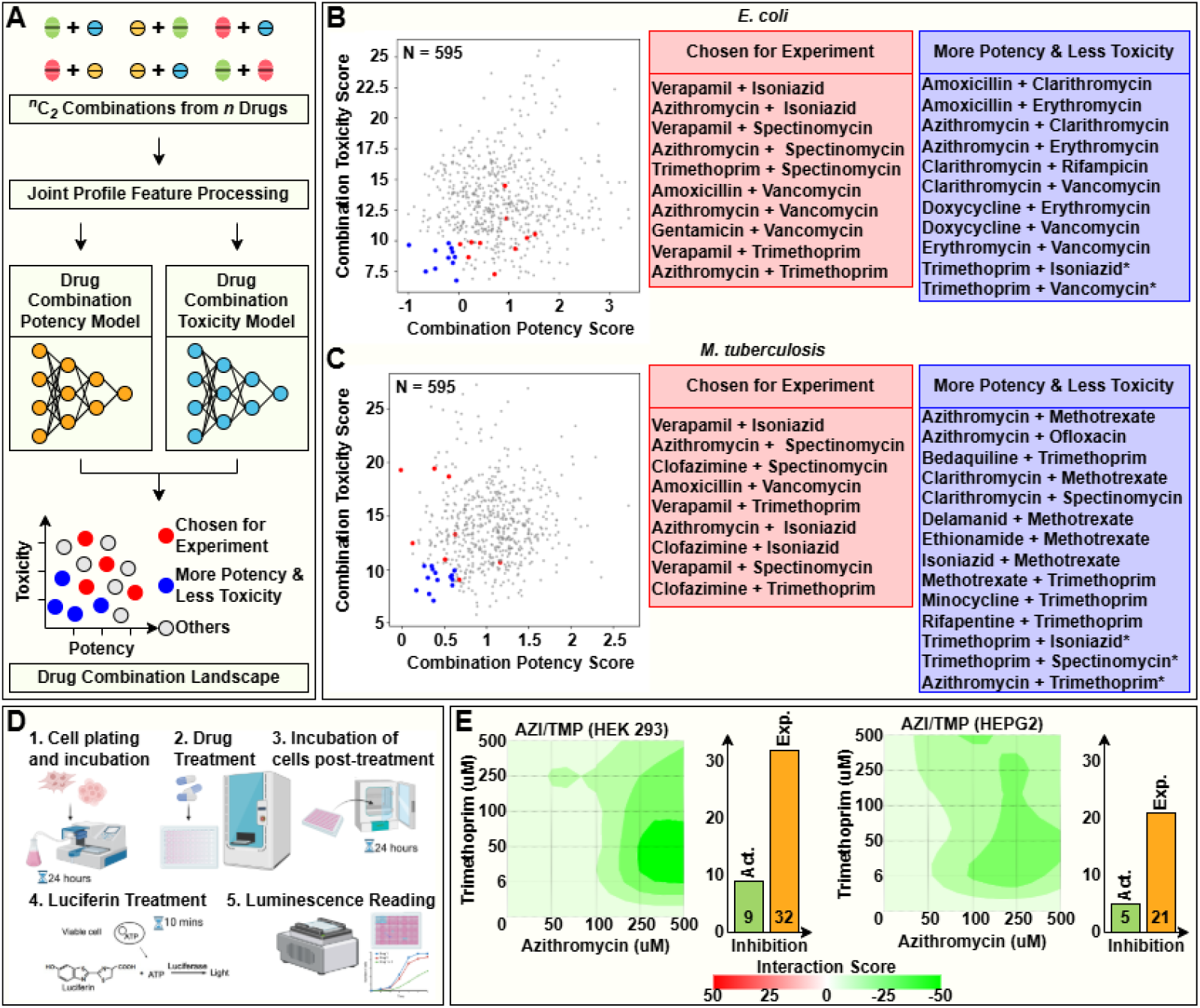
Drug Combination Landscapes for *E. coli* and *M. tuberculosis* and In Vitro Validation through Cytotoxicity Assays. **(A)** Schematic overview of generating a Drug Combination Landscape using both potency and toxicity models. Drug combinations with high potency and low toxicity are indicated in blue. Those selected for experimental validation are marked in red, and those meeting both criteria are denoted with an asterisk (*). **(B)** Drug Combination Landscape for *E. coli*, showing 595 pairwise combinations derived from 35 drugs. **(C)** Drug Combination Landscape for *M. tuberculosis*, also depicting 595 pairwise combinations from 35 drugs. **(D)** Experimental setup for in vitro combination cytotoxicity assays, utilizing the CellTiter-Glo Cell Viability Assay to measure luminescence-based cell viability in HEK293 (Kidney) and HEPG2 (Liver) cells. **(E)** Heatmaps illustrating interaction (or synergy) scores using Loewe scoring metric at various dose levels for the Azithromycin and Trimethoprim combination, noted for its highest negative synergy scores (Supplementary Table 8). Bar charts visualize expected and actual inhibition at optimal drug concentrations showing the lowest interaction scores in kidney (AZI: 250 µM, TMP: 50 µM) and liver (AZI: 100 µM, TMP: 6 µM) cells.

### *In vitro* Experimental Evaluation of Drug Combinations for Toxicity in Human Kidney and Liver Cell Lines and potency in *E. coli*

We selected fifteen drug combinations for experimental validation of model predictions of combination drug toxicity in human cell lines *in vitro*. Combinations were selected to include a range of potency and toxicity outcomes and included combinations with clinical relevance in both *E. coli* and *M. tuberculosis* infections. To increase the variety of combinatorial mechanisms, selected antibiotics spanned multiple drug classes, including aminoglycosides, macrolides, and folic acid synthesis inhibitors.

The drug Vancomycin is commonly used to treat gram positive infections and is generally considered ineffective against gram-negative bacteria due to its inability to penetrate bacterial cell membranes [30]. Further, it is known to cause renal toxicity in approximately 25% of patients [31]. Despite its known limitations, CALMA predicted specific Vancomycin combinations with macrolide antibiotics to be both potent against *E. coli* and relatively safer for human cells than Vancomycin alone, prompting us to test these combinations through *in vitro* experiments (Supplementary Figure 3A). Azithromycin, Clarithromycin, and Erythromycin are macrolide antibiotics that inhibit bacterial protein synthesis. CALMA predicted combinations with these drugs to be safe and potent against both *E. coli* and *M. tuberculosis*. We selected Azithromycin combinations for experimental testing due to Azithromycin’s superior treatment span and tolerance compared to these other macrolide antibiotics [32], [33].

The majority of combinations predicted to be potent and less toxic against *M. tuberculosis* involve Trimethoprim or Methotrexate, both of which modulate folic acid metabolism (Figure 2C). Isoniazid, a first-line antibiotic for treating tuberculosis caused by *M. tuberculosis*, is well-documented for causing acute clinically apparent liver injury [34]. We therefore also tested Isoniazid combinations through in vitro experiments. Given the reported renal toxicity of Vancomycin and liver toxicity of Isoniazid, we selected human kidney (HEK293) and human liver (HEPG2) cell lines for combinatorial cytotoxicity screening using the CellTiter-Glo assay (Methods; Figure 2D). CALMA also predicted several potentially promising antituberculosis combinations involving repurposed drugs with reduced toxicity in human cells, which were selected for testing. This includes combinations with Verapamil, a non-antibiotic drug, is used for treating high blood pressure and preventing chest pain, Clofazimine, an antimycobacterial drug indicated for leprosy treatment, and Spectinomycin, an antibiotic used for treating gonorrhea (Supplementary Figure 3B).

The experimental data were quantified using the tool SynergyFinder+ [35], which computes the Synergy Score—quantifying the activity of drug combinations relative to individual drugs (refer to Methods). As the name suggests, the Synergy Score measures the synergistic effect of the combination. For toxicity evaluations in human cell lines, we focus on lower Synergy Scores, indicating that the drug combinations exert less cytotoxicity compared to individual treatments. Supplementary Table 8 lists the different combinations and their corresponding Synergy Scores in both HEK293 and HEPG2 cells. We tested a broad dose range up to 500uM as antibiotics are known to accumulate at high concentrations in these organs [36].

Our experimental analysis identified three promising combinations with the highest antagonism (i.e. reduced toxicity): Azithromycin & Vancomycin, Isoniazid & Trimethoprim, Azithromycin & Trimethoprim (Supplementary Figures 5 and 6, Supplementary Table 8). The Azithromycin and Trimethoprim combination shows the highest negative synergy scores, i.e., lower cell inhibition than expected at all drug concentrations tested (Figure 2E). The Isoniazid and Trimethoprim combination is also predicted to be synergistic by CALMA, and has shown reduced toxicity in both Kidney and Liver cells. The Azithromycin and Vancomycin combination shows strong negative interaction scores (indicating less toxicity) at higher Azithromycin concentrations. Interestingly, the Trimethoprim and Vancomycin combination shows positive interaction scores (indicating higher toxicity) in Kidney cells but not in Liver cells, indicating the increase in renal toxicity. Additionally, combinations of Azithromycin & Vancomycin, Azithromycin & Trimethoprim, and Vancomycin & Trimethoprim were further evaluated for cytotoxicity in HK2 (kidney) and HEP3B (liver) cell lines to ensure the findings were robust to variability between cell types (Supplementary Table 14, Supplementary Figure 10). The results supported our previous findings, with combinations of Azithromycin & Vancomycin or Trimethoprim resulting in a strong antagonistic score, with reduced cytoxicity than expected. Notably, none of these combinations exhibited antagonism when tested against *E. coli* MG1655 strain; thus these combinations show reduced toxicity in human cells but not in *E. coli* (Supplementary Table 13, Supplementary Figure 9).

### Interpreting CALMA using Model Weights and Feature Knock-off Analyses

The use of metabolism-inspired ANNs in CALMA offers a dual advantage: (i) the non-linear transformations in the ANN help uncover hidden patterns in the data that traditional machine learning approaches, such as Random Forests, may miss; and (ii) the integration of GEMs with the ANN provides a framework for generating interpretable and insightful analyses of the mechanisms governing the potency and toxicity of drug combinations at the molecular level. Two methodologies, weight analysis, and feature knock-off analysis, were employed to interpret the ANN model described below (Figure 3A).

**Figure 3.**
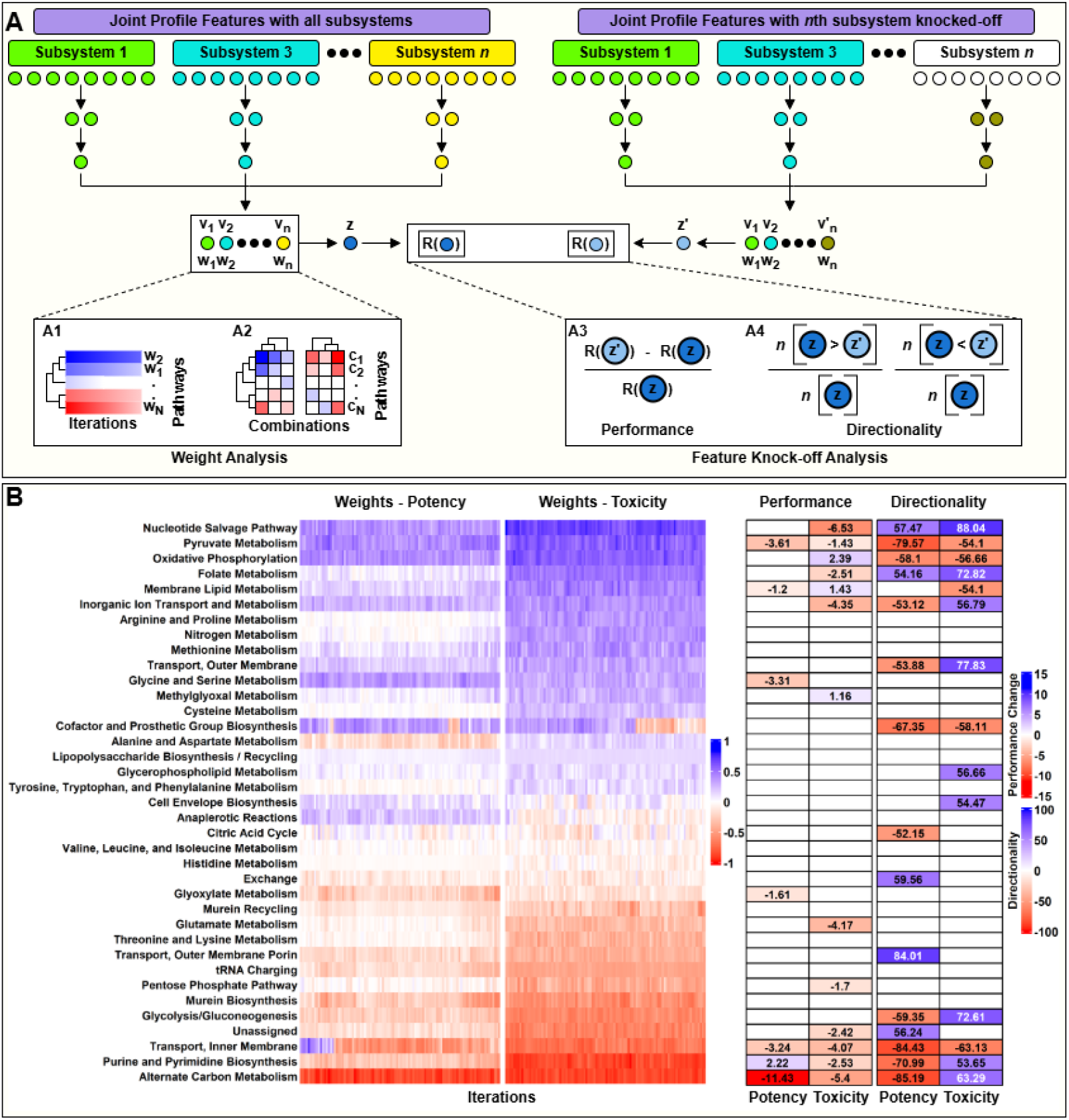
Model Interpretation using Weight and Feature Knock-off Analyses with Pathway Contributions to Potency and Toxicity Predictions. **(A)** Schematic representation of model interpretation methods: **Weight Analysis** involves (A1) examining the weights assigned to individual pathways by trained models for predicting potency and toxicity, and (A2) assessing contributions of pathways to the top and bottom 20 potent and toxic combinations. **Feature Knock-off Analysis** includes (A3) comparing model performance, and (A4) assessing prediction directionality with and without specific pathway features. **(B)** Heatmap visualizes normalized weights for different pathways in *E. coli* models for drug combination potency and toxicity, showing the effect of removing pathway-specific features on model performance (note that changes within −1% to 1% are not considered significant and are omitted) and prediction directionality (note that changes within −50% to 50% are also omitted).

We analyzed the neurons’ weights in the second hidden layer as indicators of the importance assigned to different pathways by the ANN model. The approach is illustrated in Figure 3A, with the weights for different pathways for the drug combination potency and toxicity models in *E. coli* depicted in Figure 3B. Notably, pathways such as Nucleotide Salvage Pathway, Oxidative Phosphorylation, and Pyruvate Metabolism show positive weights for both models, whereas Alternate Carbon Metabolism and Purine and Pyrimidine Biosynthesis exhibit negative weights for both models. Interestingly, the Folate Metabolism pathway has a higher weight in the toxicity model than in the potency model, highlighting its potential significance in drug combination toxicity. A similar heatmap comparing weights in drug combination potency and toxicity models for *M. tuberculosis* is shown in Supplementary Figure 4A. However, this analysis does not provide a quantitative impact of pathways on the final prediction score as the weights do not necessarily correlate with the direction of contributions from those neurons to the final output prediction.

To quantitatively assess the impact of each pathway on the final prediction score, we analyzed the weighted contributions of the neurons in the second hidden layer to the output potency or toxicity scores for the most and least potent or toxic combinations (Figure 3A, Methods). In addition to individual contributions, the mean contribution of each pathway to the final prediction scores for these combinations is presented as bar charts (Figure 4A). Alternate Carbon Metabolism emerges as the most important pathway influencing drug combination potency scores for *E. coli*, with a mean positive contribution (0.84) to top antagonistic combinations and a mean negative contribution (−0.38) to top synergistic combinations. Since lower potency scores indicate more synergistic combinations, Alternate Carbon Metabolism contributes to a decrease in the final output score for synergistic combinations and an increase in antagonistic combinations. Several pathways contribute to drug combination toxicity predictions (Figure 4A), including Purine and Pyrimidine Biosynthesis, Alternate Carbon Metabolism, Transport (Inner Membrane), and Nucleotide Salvage Pathways. With lower toxicity scores indicating less toxic combinations, the Nucleotide Salvage Pathway, in particular, shows the highest negative mean contribution (−0.28) to less toxic combinations and a highly positive mean contribution (1.31) to more toxic combinations. While other pathways like Purine and Pyrimidine Biosynthesis and Alternate Carbon Metabolism also contribute positively to toxicity scores, they do not exhibit the same degree of differentiation between less and more toxic combinations. Transport (Inner Membrane) contributes negatively to less toxic combinations and positively to more toxic combinations but with less impact than the Nucleotide Salvage Pathway. A similar heatmap of the weighted contributions of pathways for *M. tuberculosis* is shown in Supplementary Figure 4B.

**Figure 4.**
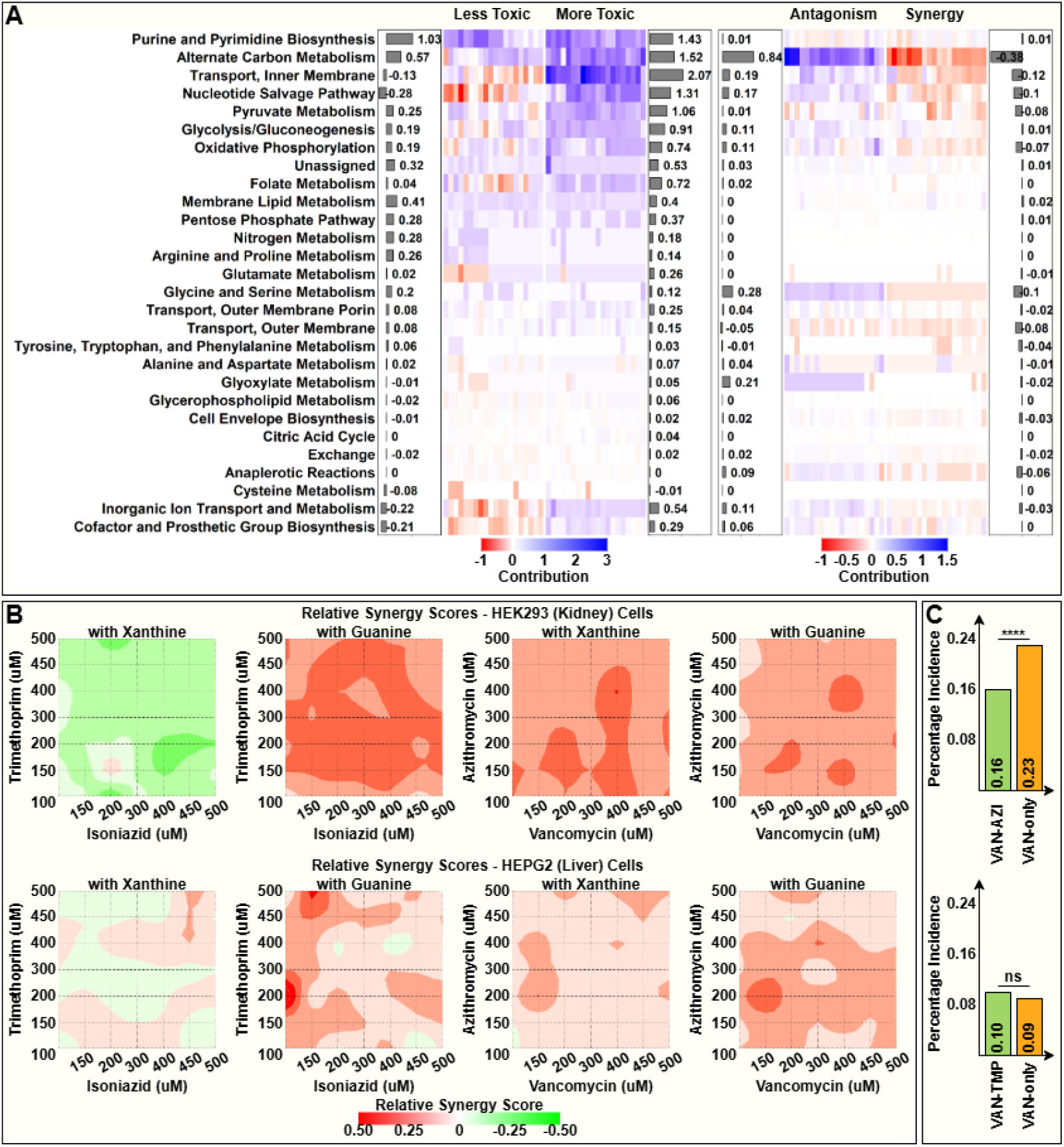
Analysis of Pathway Contributions, Synergy Scores, and Drug Combination Efficacy. **(A)** Cluster heatmap depicting the weighted contributions of various pathways to the top and bottom 20 combinations in terms of potency and toxicity for *E. coli*. Accompanying bar charts show the mean contributions of each pathway to the final prediction scores. **(B)** Heatmaps illustrating relative synergy scores of selected drug combinations in the presence of nucleotide metabolites, with positive scores (red) indicating decreased cell viability (or increased cell inhibition) when nucleotide metabolites are present. **(C)** Bar chart comparing the percentage incidence of Nephrotoxicity with the combination of Vancomycin with Azithromycin and Vancomycin with Trimethoprim with the Vancomycin-alone group based on mining of patient health records (^****^p<0.0001, ns: not significant).

In addition to weight analysis, feature knock-off analysis provides insights into the importance of specific features or sets of features by assessing the model’s performance and predictions in their presence and absence. The absence of a particular subsystem is simulated by setting the corresponding features to zero and assessing the relative change in model performance and relative change in potency/toxicity scores predicted by the model. This analysis also indicates that Alternate Carbon Metabolism is the most crucial pathway for drug combination potency, correlating with the highest negative change in model performance (−11.43%). In contrast, the Nucleotide Salvage Pathway is the most significant for drug combination toxicity, with a notable reduction in model performance (−6.53%).

### *In vitro* Evaluation of the Role of the Nucleotide Salvage Pathway in Drug Combination Toxicity

From the interpretative analyses, we determined that the Nucleotide Salvage Pathway is the most influential pathway governing drug combination toxicity. To further corroborate the computational prediction of this pathway’s importance, we designed in vitro experiments involving drug combination toxicity screening with nucleotide supplementation. We aimed to determine whether toxicity was decreased when an abundance of nucleotides disincentivized cells from utilizing the Nucleotide Salvage Pathway, and conversely, whether depleting cellular dNTP levels would result in enrichment of nucleotide salvage. HEK293 (Kidney) and HEPG2 (Liver) cells were treated with various metabolites and inhibitors related to nucleotide synthesis and salvage. The metabolites employed for these experiments included Guanine (a purine nucleotide), Thymine (a pyrimidine nucleotide), and Xanthine (a purine analog). Additionally, Gemcitabine (a nucleoside inhibitor) was selected to test perturbance of the nucleotide salvage pathway. We selected three notable combinations from previous toxicity experiments (Azithromycin & Vancomycin, Isoniazid & Trimethoprim, Trimethoprim & Vancomycin). We observed the effects of these drug combination treatments on human cell lines in the presence and absence of these metabolites.

The experimental output from the assay was processed to calculate Bliss synergy scores for drug combinations both in the presence and absence of these metabolites (Methods, Supplementary Figures 7 and 8, Supplementary Tables 9 and 10). In most cases, the addition of these metabolites increased the Bliss synergy scores, indicating that the drug combination treatments became more synergistic and, consequently, more toxic. Interestingly, the combination of Isoniazid and Trimethoprim, which was found to be safe for both kidney and liver cells, exhibited a decrease in synergy score in the presence of Xanthine and Thymine in HEK293 cells and the presence of Thymine in HEPG2 cells. Moreover, the combination of Azithromycin and Vancomycin, which was observed to reverse the renal toxicity of Vancomycin, showed reduced cell viability in the presence of Guanine, Xanthine or Gemcitabine in HEK293 cells (Figure 4B).

### Mining of health records corroborates reduced nephrotoxicity of vancomycin combinations identified by CALMA

To obtain real-world evidence for some of the top combinations prioritized by CALMA, we mined health records of over 330 million deidentified US patients that were part of the Komodo Healthcare Map™. We identified patients who had a reported use of Vancomycin, and further identified sub-populations who also had co-administered Trimethoprim / Azithromycin within 5 days of the first vancomycin use. Filtering for patients who had Vancomycin co-administered with either of these antibiotics would likely also enrich a population with more comorbidities. Thus, to account for such potential biases, a propensity score matching analysis was performed. We first calculated a propensity score for each patient in the population using various comorbidities and medical history factors. Then, for each of the 182,565 patients who were identified with co-administration of Vancomycin and Azithromycin, we identified a patient with the most similar propensity score from the Vancomycin only group.

When we measured the occurrence of a nephrotoxicity code in the 6 months after Vancomycin in these populations, we noticed a marked decrease in incidence with Azithromycin (291 cases out of 182,565 in Vancomycin + Azithromycin group, or 0.16%, vs 416 cases out of 182,565 in the Vancomycin only group, or 0.23%; t-test p-value = 2.53E-06). However, a similar analysis with Trimethoprim co-administration did not yield any changes in incidence rates (361 vs 396 cases out of 402,949 for Vancomycin-only vs Vancomycin+Trimethoprim; t-test p-value = 0.203) (Figure 4C). This is consistent with the *in vitro* toxicity assays that revealed antagonism of Vancomycin with Azithromycin but not with Trimethoprim. These results suggests that Azithromycin may reduce the renal toxicity of Vancomycin. While the nephrotoxicity incidence is low, given the large number of patients taking these drugs (3,877,685 patient cases in the Komodo dataset reported use of Vancomycin in 2016-2025), these small percentages translate to thousands of cases of nephrotoxicity.

## DISCUSSION

In this study, we introduce CALMA, a computational tool designed to assist in the development of potent and safe antimicrobial drug combinations. CALMA provides several advantages over existing approaches. First, it integrates two distinct computational frameworks in biology and computer science: Genome-scale Metabolic Models (GEMs) and Artificial Neural Networks (ANNs). The metabolic pathway information from GEMs is utilized to design the ANN model’s architecture, providing a knowledge bias that aids in efficient training and model development. Second, CALMA not only predicts drug combination potency scores but also predicts drug combination toxicity scores, ensuring both the efficacy and safety of combination therapies. To our knowledge, no existing framework predicts both potency and toxicity scores for drug combinations. Third, the development of drug combination potency and toxicity models facilitates the generation of a drug combination landscape, optimizing the potency and safety of drug combinations and narrowing the search space. Fourth, the metabolism-inspired design of the ANN model provides insights into the mechanisms of action governing drug combinations at the pathway level. Deep learning models such as ANNs often face criticism for their black-box nature and lack of interpretability despite their high performance. From the model interpretation analyses (Figures 3B and 4A), we inferred that Alternate Carbon Metabolism is a key pathway governing drug combination potency, whereas the Nucleotide Salvage Pathway is likely crucial for drug combination toxicity. Further examination revealed that adding nucleotide metabolites to drug combination treatments increases cell inhibition in human cells.

A major contribution of this work is the generation of the drug combination landscape. By narrowing down the list of potential combinations, we selected several combinations for testing their safety in human cells using cell viability assays (Figure 2D). Specifically, our experimental analysis (Supplementary Table 8) identified three promising combinations with reduced toxicity in multiple human kidney and liver cell types as compared to individual drugs: Azithromycin & Trimethoprim, Azithromycin & Vancomycin, and Isoniazid & Trimethoprim. Isoniazid is a first-line treatment for *M. tuberculosis* and inhibits mycolic acid synthesis, an essential component of mycobacterial cell walls. Azithromycin inhibits bacterial protein synthesis and also exhibits immunomodulatory effects, while Trimethoprim inhibits the enzymatic pathway involved in the synthesis of tetrahydro-folic acid. Studies have shown that combining Trimethoprim, Sulfamethoxazole (a folate inhibitor) with Isoniazid has a bactericidal effect on *M. tuberculosis* and prevents resistance [37]. Our *in vitro* experiments revealed that the Trimethoprim and Isoniazid combination also has antagonistic effects on human cells and thus can be a promising combination for treating Tuberculosis infections with reduced hepatic side-effects, leading to better patient adherence to the full course of treatment. The Azithromycin & Vancomycin combination was found to have antagonistic effects on kidney cells and showed mild synergy against *E. coli*. This trend also held up in an analysis of health records of patients, wherein patients had lower chance of nephrotoxicity when taking these drugs in combination compared to taking vancomycin alone. This suggests Azithromycin may mitigate the renal toxicity of Vancomycin.

While CALMA aids in designing safe and effective combinations, it has limitations. ANN models require large datasets for optimal performance. The scarcity of high-quality drug combination interaction data poses a challenge for training ANN models, as seen in the benchmarking analysis. Additionally, GEMs depend on experimentally generated omics data under different drug conditions to simulate metabolic reaction fluxes. Thus, CALMA cannot predict outcomes for drugs lacking omics data, which could be overcome by approaches that computationally predict drug primary and secondary targets [38], [39]. We anticipate that CALMA will become a valuable instrument in designing tailored combination therapies, contributing to global efforts to combat drug resistance. Beyond bacterial infections, CALMA has the potential to be extended to other microbial species, such as fungi, and to other disease areas where drug resistance is a substantial concern, such as cancer.

## METHODS

### ANN Model Development and Evaluation

The schematic for model development is detailed in Supplementary Figure 1. Below, we outline the steps involved in model training, validation, and testing:

Step 1: The dataset is divided into two parts: a training set and a test set. The training set is used for model training and validation, while the test set is reserved for model evaluation.

Step 2: The training set is further divided into two subsets: the training subset and the validation subset. The training subset is used specifically to train the model, and the validation subset is employed to validate the model and select optimal parameters prior to making final predictions on the test dataset.

Step 3: The training subset is utilized to train the model for 250 epochs using hyperparameters selected through hyperparameter tuning (refer to Methods).

Step 4: At each epoch, predictions are generated using the validation subset from Step 2.

Step 5: Spearman rank correlations are computed for each epoch’s predictions on the validation subset.

Step 6: Based on the Spearman rank correlation values for each epoch, the top ten epochs with the highest correlation scores are selected.

Step 7: Using the trained model from Step 3 and the top ten epochs identified in Step 6, predictions are made on the test dataset. The final test predictions are the average of the predictions from these ten epochs.

Step 8: The test predictions from Step 7 and the test dataset from Step 1 are used to compute the final Spearman rank correlation score for the test set.

### Simulation of Metabolic Reaction Fluxes Using GEMs for *E. coli* and *M. tuberculosis*

We utilized the *E. coli* GEM model iJO1366 and the *M. tuberculosis* GEM model iEK1008 to simulate metabolic reaction fluxes under steady-state conditions. For the simulation of the objective function, additional constraints were applied to the models using chemogenomics data and transcriptomics data for *E. coli* and *M. tuberculosis* GEMs, respectively. Specifically, information on differential gene regulation was incorporated as constraints for the GEMs.

Chemogenomics data measure single-gene knock-out (KO) fitness, where a high fitness score indicates that the gene is non-essential, while a low fitness score signifies that the gene is essential for bacterial growth. Genes were further classified as up- and down-regulated based on KO fitness score thresholds of 2 and −2, respectively.

Transcriptomics data, on the other hand, provide information on gene expression levels under both drug-treated and untreated conditions. Similarly to chemogenomics data, expression values were classified as up- and down-regulated based on thresholds of 2 and −2, respectively. These individual sets of differentially regulated genes were integrated into their corresponding GEMs using a linear optimization version of the integrative metabolic analysis tool (iMAT).

Moreover, employing the linear iMAT algorithm [40]–[42] necessitates the fine-tuning of three constraint-based modeling parameters: kappa, rho, and epsilon [43]. Kappa and rho assign relative weights to “off” and “on” reactions associated with their differentially regulated (or expressed) genes, respectively, in their contribution to the objective function (e.g., biomass). Epsilon determines the minimum flux through “on” reactions. We utilized the same parameter values for both *E. coli* and *M. tuberculosis* GEMs as those used in the recently introduced CARAMeL approach [9] for predicting drug combination outcomes using GEMs. The parameter values for kappa, rho, and epsilon for the iJO1366 GEM (*E. coli*) are 0.01, 0.01, and 1, respectively, while for the iEK1008 GEM (*M. tuberculosis*), the values are 0.01, 0.01, and 0.01, respectively.

### Generation of Sigma and Delta Scores from Reaction Flux Profiles

Reaction flux profiles generated after the simulation of GEMs correspond to individual drug treatments. These reaction flux profiles are binarized based on differential flux activity (either positive or negative) relative to the baseline reaction flux. Upon binarization, each reaction in the profile of an individual condition is represented by a set of two binary numbers, *V*_*positive*_, and *V*_*negative*_ depending on whether the reaction exhibits positive or negative differential flux activity.

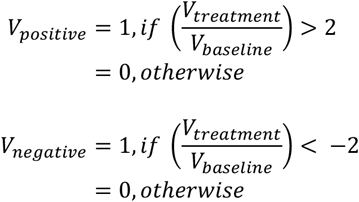

The binarized profiles of conditions are subsequently used to calculate sigma and delta scores for drug combinations. Sigma scores represent the combined effect of the drugs in a combination, whereas delta scores represent the unique effect of each drug in the combination [7], [9]. The sigma and delta scores are computed mathematically as follows:

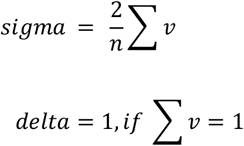

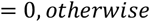

Where, *v* is the binary reaction flux, and *n* is the number of drugs in a combination. Given that each reaction is represented by a set of two binary numbers, the sigma and delta scores are derived from these binary values. Consequently, a single reaction is characterized by four features within the joint profile features.

### Hyperparameter Tuning for ANN Model Optimization

The ANN model is characterized by two sets of parameters: weights and hyperparameters. The model weights are learned during the training phase, whereas hyperparameters must be specified before model training. Various hyperparameters, such as the number of hidden layers, the number of neurons in each hidden layer, loss function, optimizer, and learning rate, can significantly affect the model’s performance.

Hyperparameter tuning is the process of optimally selecting hyperparameters. This process involves training ANN models with different sets of hyperparameters and comparing their performance to choose the optimal configuration. The GEM directly informs the determination of the number of hidden layers and the number of neurons within those hidden layers. For other hyperparameters, such as the loss function, optimizer, and learning rate, hyperparameter tuning is conducted. Detailed information on the various hyperparameters and their corresponding options used for tuning is provided in Supplementary Table 1.

Both ANN models derived from *E. coli* and *M. tuberculosis* GEMs are subjected to the hyperparameter tuning process. We trained the ANN models under different hyperparameter settings using five-fold cross-validation to predict drug combination potency scores. Spearman rank correlation was employed to evaluate model performance for each set of hyperparameters. The performance metrics from the *E. coli* and *M. tuberculosis* GEM-derived ANN models were then averaged. The rationale behind averaging the performance metrics from both models is to ensure a consistent set of hyperparameters for both. Performance metrics (i.e., rank correlation) for the hyperparameter tuning process are presented in Supplementary Table 2. The selected hyperparameters—loss function (mean absolute error), learning rate (0.1), and optimizer (stochastic gradient descent)—were used to train all the ANN models in this study.

### Comparison of CALMA potency prediction with literature

For the first case, we utilized the dataset from the INDIGO study, which includes 171 pairwise drug combination outcomes among 19 drugs [7]. We employed a five-fold cross-validation (CV) approach for benchmarking. CALMA achieved a highly significant correlation score between experimental scores and predicted outputs (R = 0.56, p ≈ 10^-14; see Supplementary Figure 2A).

In the second case, we leveraged the dataset from the MAGENTA study [28], consisting of 282 interactions, of which 171 were derived from the INDIGO study. Twenty-eight interactions were measured in both LB and M9 glucose media, while the remaining 55 interactions were measured in M9 glycerol media. The interactions derived from INDIGO, as well as those measured in both LB and M9 glucose media, were used as the training set, while interactions measured in M9 glycerol media served as the test set. CALMA provided similar accurate predictions comparable to MAGENTA (R = 0.55, p ≈ 10^-6; see Supplementary Figure 2B).

For the third case, INDIGO-MTB study [8] measured 232 multi-drug interactions, ranging from two-way to four-way interactions. Similar to the INDIGO approach, a five-fold CV method was used for benchmarking. CALMA yielded highly significant and accurate predictions (R = 0.44, p ≈ 10^-13; see Supplementary Figure 2C).

In the fourth case, we retrained the model using the dataset from INDIGO-MTB to predict drug combination outcomes for 57 multi-drug TB regimens used in randomized clinical trials. The efficacy outcomes for this dataset were determined based on sputum conversion rates after two months of treatment. We observed that CALMA’s predictions significantly correlated with sputum conversion rates (R = 0.42, p ≈ 10^-4; see Supplementary Figure 2D).

### Toxicity Assessment and Calculation of Proportional Reporting Ratio (PRR) Using the TWOSIDES Database

The FDA Adverse Event Reporting System (FAERS) is a database that provides information on side effects and adverse reactions caused by various treatments (single-drug or multi-drug). TWOSIDES is a comprehensive dataset derived from FAERS, offering information on drug combination-side effect relationships. It encompasses over 3,300 drugs and 63,000 combinations linked to millions of potential adverse reactions.

TWOSIDES details the various side effects observed from specific drug combination treatments and quantifies the toxicity of each combination via a contingency table (as shown in Figure 1F). For each side effect associated with a drug combination treatment, TWOSIDES provides the following information, where A, B, C, and D are the number of cases corresponding to a different setting.

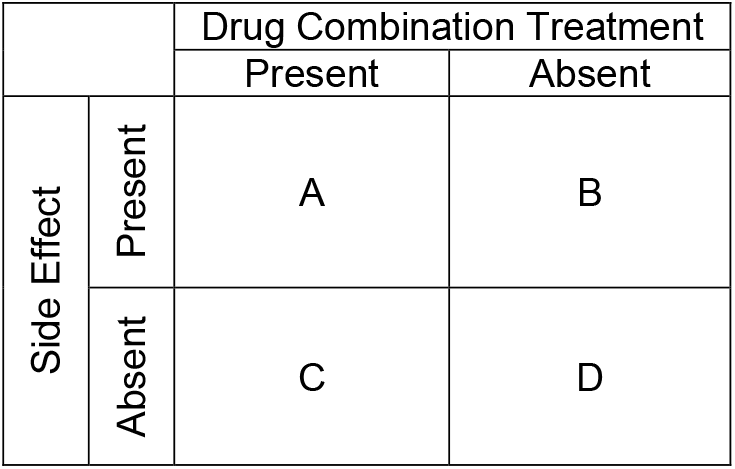

|  |  | Drug Combination Treatment |  |
| --- | --- | --- | --- |
|  |  | Present | Absent |
| Side Effect | Present | A | B |
|  | Absent | C | D |

Using the information, the Proportional Reporting Ratio (PRR) is calculated according to the following mathematical formula.

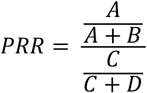

Essentially, the PRR score is the ratio of the fraction of cases where a side effect was observed in the presence of the drug combination treatment to the fraction of cases where the side effect was observed in the absence of the drug combination treatment. Based on this definition, it is hypothesized that the PRR score measures relative toxicity. A higher PRR score indicates greater toxicity.

In addition to providing the PRR score, the database also includes the standard error associated with the estimation (or calculation) of the PRR score itself. The Error-PRR is calculated using the following mathematical formula.

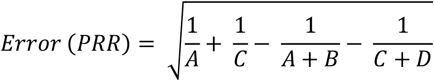

Furthermore, the significance of the occurrence of a particular side effect due to a specific drug combination treatment is determined by the following condition [44].

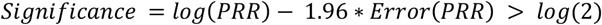

To ensure the inclusion of relevant drug combination-side effect pairs, we used the significance condition to filter the data. In cases where a particular drug combination is associated with multiple side effects, cumulative values for A, B, C, and D were calculated to determine a single PRR score for each combination. These cumulative values were obtained by summing the corresponding values for each significant side effect, followed by computing the PRR score using the formula provided above.

### Generation of Drug Combination Landscape based on Potency and Toxicity Predictions

The drug combination potency and toxicity models for both *E. coli* and *M. tuberculosis* were employed to generate predictions for 595 pairwise combinations among 35 individual drugs. Detailed information on the 35 drugs used for both *E. coli* and *M. tuberculosis* is provided in Supplementary Tables 4 and 5. The drug combination potency prediction model is trained on scoring metrics such as Loewe or Bliss, which quantify the effect of a combination compared to individual drugs. Conventionally, lower scores indicate a more potent (or synergistic) combination. Thus, when analysing the drug combination landscape, we focus on the left section of the x-axis. Similarly, the drug combination toxicity prediction model utilizes PRR scores. As illustrated in Figure 1F, higher PRR scores suggest greater toxicity of the combination, directing our interest towards the bottom section of the y-axis. To filter out the most and least toxic and potent combinations, a threshold of 15% was utilized. We defined the following categories of combinations.

| Category | Combinations |
| --- | --- |
| more_potent | Lowest 15% potency scores |
| less_potent | Highest 15% potency scores |
| more_toxic | Highest 15% toxicity scores |
| less_toxic | Lowest 15% toxicity scores |

Using the definition of these categories, we also define the following category of combinations.

| Category | Condition |
| --- | --- |
| More Potent & Less Toxic | more_potent ∩ less_toxic |
| More Potent & More Toxic | more_potent ∩ more_toxic |
| Less Potent & Less Toxic | less_potent ∩ less_toxic |
| Less Potent & More Toxic | less_potent ∩ more_toxic |

where, A *∩* B represents the list of combinations present in both lists A and B.

### Combination Cytotoxicity Screening in HEK293 and HEPG2 Cells

#### Mammalian Cell Culture and Maintenance

The HEK-293, HEPG2, HK2, and HEP3B cell lines were obtained from the American Type Culture Collection (ATCC, USA). Cells were cultured under standard conditions and stored in Gibco Recovery Cell Culture Freezing Medium at 2 × 10^6^ cells per aliquot. For routine culturing and maintenance, cells were grown in Dulbecco’s Modified Eagle Medium (DMEM, Life Technologies) supplemented with 10% Fetal Bovine Serum (FBS) and 1% penicillin/streptomycin (Pen/Strep). Cultures were maintained at 37°C with 5% CO^2^.

#### Bacterial Strains, Compounds, and Reagents

We obtained the *E. coli* MG1655 strain used in this work from the USDA-ARS Culture Collection (NRRL). The bacterial strain was selected due to being well-represented in our antimicrobial interaction training dataset, as well as being a well-studied model organism. We purchased antibiotic powders through Cayman Chemical (Ann Arbor, USA). Antibiotic powder stocks were kept at 4°C, and liquid stocks were made in ddH_2_O or dimethyl sulfoxide (DMSO) and stored at –20°C for the duration of experimental use. The *E. coli* medium used for overnight liquid culturing and experimental assays was Luria-Bertani Broth (LB, BD Biosciences, USA) and 12 g of agar was supplemented to make solid plates.

#### Checkerboard Assays

The activity of antibiotics in combination was evaluated using a checkerboard assay. Briefly, both compounds were two-fold serially diluted in 50 μL of experimental medium within two separate flat-bottom microtiter plates starting at a concentration of 2xMIC for each compound, or the highest dose tested for drugs without measurable activity. The first compound was transferred to wells containing the second compound, so that each row contained a serial dilution of one compound while each column contained a serial dilution of the second compound in the perpendicular direction.

After the dilution process, 50 μL of medium was added to each well, followed by 50 μL of an overnight liquid bacterial culture adjusted to an optical density (OD600) of 0.08-0.1 to all wells except for the media blanks. Additionally, each plate included wells containing only a single compound, serially diluted two-fold. The plates were incubated at 37°C for 18-24 hours, and OD600 measurements were recorded. Each experiment was conducted in duplicate.

#### Cell Viability Assays

The viability and toxicity of antibiotic combinations was assessed in HEK-293, HEPG2, HK2, and HEP3B cells using the CellTiter-Glo assay (Promega, USA). Briefly, cells were seeded in 20 μL of medium at a density of 1,000 cells per well in opaque, 384-well tissue culture-treated plates and allowed to adhere overnight. After 24 hours, cells were treated with serial dilutions of single antibiotics as well as eight-concentration checkerboard combinations using a plate layout as described above. Compound dispensing was performed using the Echo Acoustic Liquid Handler (Center for Chemical Genomics, University of Michigan). Following treatment, 20 μL of fresh medium was added to each well and plates were incubated for an additional 24 hours. After incubation, 20 μL of CellTiter-Glo reagent was added to each well and, following 5 minutes of incubation at room temperature, luminescence was measured using a spectrophotometer. Results were normalized to DMSO controls, and growth curves were generated using GraphPad Prism 10.

### Quantifying Drug Combination Activity Using SynergyFinder+

To quantify the activity of drug combinations relative to individual drugs, we utilized a tool called SynergyFinder+, which employs cell viability outputs from combinatorial experimental assays to calculate a metric known as the Synergy Score. This tool provides various scoring metrics to compute the Synergy Score, with each method based on specific assumptions. Below, we describe the different scoring metrics (or methods) used to calculate the Synergy Score.

Definitions,

y1 = monotherapy effect (inhibition) of the drug 1

y2 = monotherapy effect (inhibition) of the drug 2

#### Bliss

The Bliss metric assumes that two drugs exert their effects independently. The expected combination effect is calculated based on the probability of independent events.

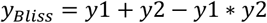

#### Loewe

The Loewe metric defines the expected effect of the combination as if a drug were combined with itself.

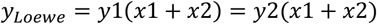

Unlike the Bliss model, the Loewe model considers the dose-response curves of individual drugs. The expected *y*_*Loewe*_ must satisfy,

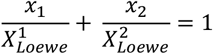

where, *x*_*1*,2_ are drug doses and 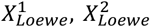 are doses of drug 1 and 2 alone that produce *y*_*Loewe*_. By utilizing 4-parameter log-logistic (4PL) curves to describe dose-response curves, the following parametric form of previous equation is derived,

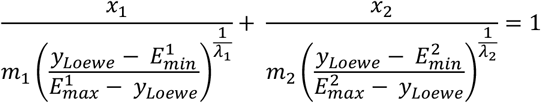

where, *E*_*min*_, *E*_*max*_ *ϵ* [0, 1] are minimal and maximal effects of the drug, *m*_*1*,2_ are the doses of drugs that produce the midpoint effect of *E*_*min*_ + *E*_*max*_, also known as relative *EC*_50_ or *IC*_50_, and *λ*_1,2_ (*λ* > 0) are the shape parameters indicating the sigmoidicity or slope of dose-response curves.

### Calculation of Bliss Synergy Scores for Nucleotide Experiments

SynergyFinder+ assists in analyzing the impact of drug combination treatments on cells compared to individual drug treatments. However, for nucleotide experiments, our objective was to analyze the effect of drug combination treatments in the presence and absence of nucleotides. Since SynergyFinder+ is not equipped to analyze these specific data points, we employed the Bliss Synergy Score methodology to compute synergy scores for drug combinations both alone and in the presence of nucleotides.

For the calculation of Bliss Synergy Scores, we defined the following parameters,

*A*_*i*_= cell viability with Drug A alone at *i* uM

*B*_*j*_= cell viability with Drug B alone at *j* uM

*V*_*N*_= cell viability with nucleotide metabolite alone at 20 uM

*V*_*i,j*_ = cell viability with both drugs A and B at *i* and *j* uM respectively

*V*_*i,j,N*_ = cell viability with drugs A and B at *i* and *j* uM resp.and nucleotide at 20 uM

Based on the assumption of the Bliss Synergy Score model, that individual components in a combination exert their effects independently, we can calculate the synergy scores as follows,

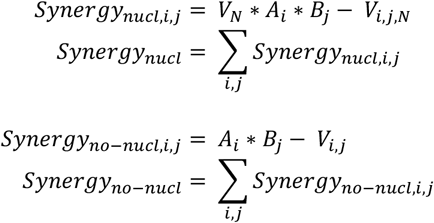

Comparing these two synergy scores provides insights into the impact of the addition of nucleotide metabolites on the efficacy of drug combination treatments in human cell lines. If *Synergy score*_*nucl*_ is greater than *Synergy score*_*no–nucl*_, it indicates that the addition of the nucleotide metabolite makes the drug combination more synergistic and, therefore, more toxic, leading to decreased cell viability.

### Interpreting CALMA ANN model using weights and contributions

ANN models learn internal parameters (or weights) from the dataset during training through backpropagation. Weights are initialized randomly and are updated as training progresses. As illustrated in Figure 1B, the value of neurons in the second hidden layer is dependent on input features from specific pathways (or subsystems). These neurons then contribute to the final model output—either drug combination potency or toxicity scores.

Let *v1, v2, ……, vN* and *w1, w2, ……, wN* represent the values and the weights of the neurons in the second hidden layer, respectively. Here, N denotes the number of neurons in the second hidden layer corresponding to the number of pathways (or subsystems) in the model.

We define *ci = vi* * *wi*, (where *i* ranges from *1 to N*) as the contribution (or weighted output) from each neuron in the second hidden layer to the final output score (potency or toxicity).

Using these definitions, the model’s final output score (*z*) is given by

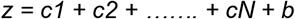

where b is the bias learned during the model training.

### Mining health records

For the retrospective cohort study, a comprehensive clinical database from Komdo Health’s Healthcare Map, named the Komodo Research Dataset (KRD) was used (data version: February 2025). This dataset includes integrated inpatient, outpatient, and pharmacy records. Patients receiving doses of Vancomycin, Azithromycin and Trimethoprim were identified from the use of the respective drug-NDC codes and J/Q/S codes within the time period of 2016 - 2025. Exposure to adjunctive therapy was defined as any record of azithromycin (or trimethoprim) occurring within a +/− 5 day window relative to the first date of vancomycin administration. This window was chosen to ensure temporal alignment between vancomycin and the adjunct therapy. Patients were subsequently categorized into two groups: those receiving vancomycin alone versus those receiving combination therapy. Nephrotoxicity was defined using ICD diagnostic codes beginning with N.14 and was considered as an event if it occurred between 1 and 180 days after the index vancomycin exposure. The lower bound of 1 day was set to avoid including events potentially unrelated to vancomycin administration, while the upper bound of 180 days captured kidney injuries plausibly linked to the treatment.

Covariate Extraction and Feature Engineering: Patient demographics and clinical characteristics were extracted from the database, including age, gender, ICU admission status during index event, and length of stay. In addition, diagnostic codes were utilized to generate flags that summarized presence of key comorbid conditions in the complete history of each patients. For this study, the following flags were calculated:

- AB_FLAG: A combined indicator for infectious and parasitic diseases (ICD-10 A00–B99).
- C_FLAG: Reflecting neoplasms (ICD-10 C00–C97).
- F_FLAG: Representing mental and behavioral disorders (ICD-10 F00–F99).
- H_FLAG: Denoting diseases of the eye and ear (ICD-10 H00–H95).
- J_FLAG: Indicating respiratory system diseases (ICD-10 J00–J99).
- L_FLAG: Capturing disorders of the skin and subcutaneous tissue (ICD-10 L00–L99).
- M_FLAG: Representing musculoskeletal and connective tissue disorders (ICD-10 M00– M99)

Additionally, a pneumonia flag, indicating the presence of pneumonia, defined by relevant ICD codes recorded within a ±180-day window of the index event, were included.

Propensity Score Estimation and Stratified Matching: To address confounding from non-random treatment allocation, a propensity score was estimated using logistic regression. The dependent variable was the receipt of adjunctive therapy (combination treatment) versus vancomycin alone, and the covariates in the model comprised ICU admission status, pneumonia flag, and the seven diagnostic flags. Patients were then stratified by age (5 bins) and gender to further enhance comparability. Within each age–gender stratum, a 1:1 nearest-neighbor matching algorithm was applied using Euclidean distance on the propensity score scale. This stratified approach ensured that matched pairs were similar not only on the clinical covariates but also on key demographic characteristics.

Covariate Balance and Outcome Analysis: Post-matching, covariate balance was assessed using graphical methods (e.g., kernel density plots) and by calculating absolute differences between matched pairs. The primary outcome—nephrotoxicity—was then compared between treatment groups by computing the proportion of patients who experienced a nephrotoxic event within the 1–180 day window. Statistical significance was evaluated using t-tests.

### Additional description of top combinations

#### Azithromycin and Trimethoprim

This combination has a high negative synergy score for both kidney and liver cells, indicating antagonistic effects. The combination of Azithromycin and Trimethoprim showed weak synergistic interactions at specific concentrations when tested against *E. coli* (Supplementary Table 13, Supplementary Figure 9).

#### Isoniazid and Trimethoprim

This combination is predicted to be synergistic for both *E. coli* and *M. tuberculosis*. It is among the top 15% of safe and effective combinations for both organisms (Figures 2B and 2C).

#### Trimethoprim and Vancomycin

Vancomycin inhibits the production of peptidoglycans, essential for bacterial cell walls, particularly in Gram-positive bacteria. CALMA predicted that its combination with Trimethoprim is synergistic against *E. coli* and lies within the top 15% of combinations (Figure 2B). We found potent synergy *in vitro* between Vancomycin and Trimethoprim against *E. coli* consistent with CALMA predictions (Supplementary Table 13, Supplementary Figure 9). Our *in vitro* toxicity assay results indicated that the Trimethoprim and Vancomycin combination has a positive synergy score for kidney cells and a negative synergy score for liver cells. This observation is consistent with the renal toxicity of Vancomycin.

#### Azithromycin and Vancomycin

This combination has a negative synergy score, indicating antagonistic effects on both kidney and liver cells. This combination also showed mild synergy against *E. coli* at a Vancomycin concentration of 32 µg/mL and an Azithromycin concentration of 4 µg/mL. This suggests Azithromycin may mitigate the renal toxicity of Vancomycin.

## Data Availability

All data produced in the present study are available upon reasonable request to the authors. This paper analyzes data from the Komodo Health Platform (2016-2025) after signing a data-sharing agreement.

## Acknowledgments

This work was supported by faculty start-up funds from the University of Michigan (UM), R01AI150826 from National Institute of Allergy and Infectious Diseases, R35GM137795 from National Institute of General Medical Sciences, UM Endowment for Basic Sciences Accelerator Award, UM Research Scouts Award to S.C.

## SUPPLEMENTARY FIGURES

**Supplementary Figure 1.**
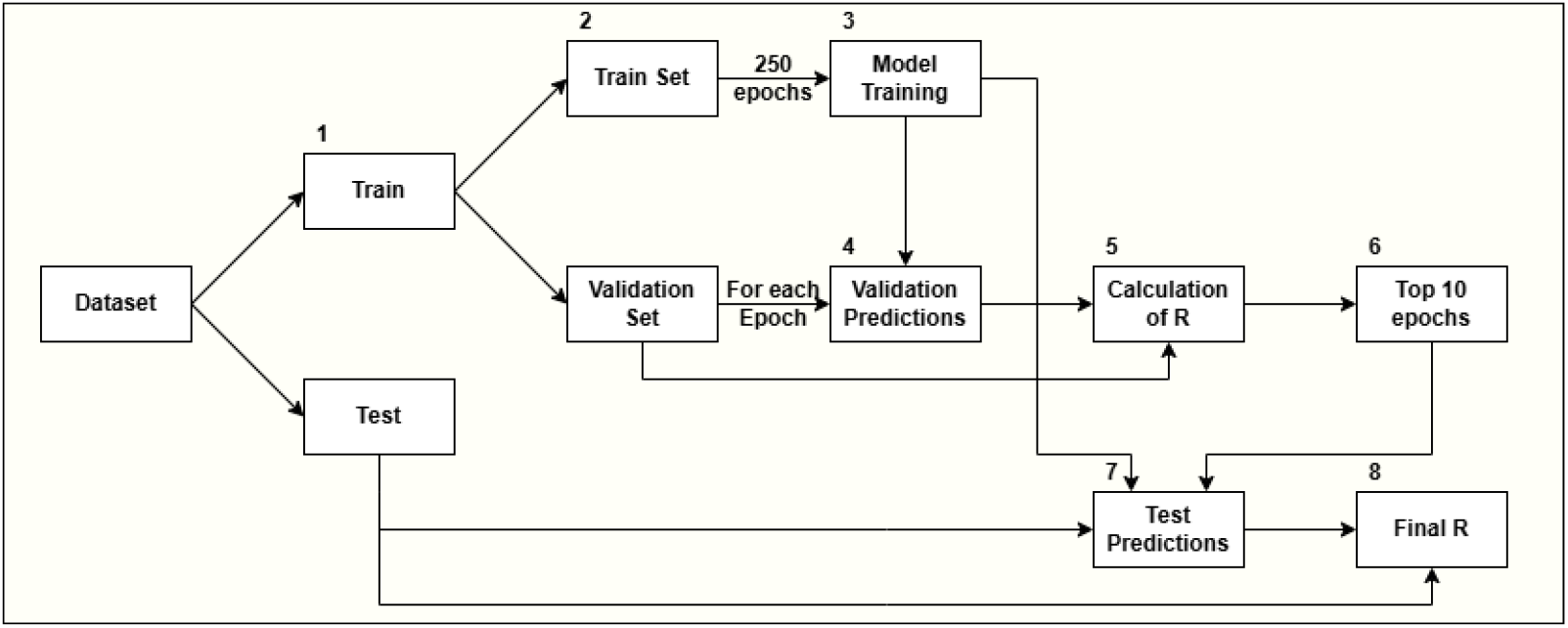
Schematic overview of the ANN model development process, detailing the use of the training set to train the model, and the use of the validation set to select the best model weights and parameters for making predictions on the test set.

**Supplementary Figure 2.**
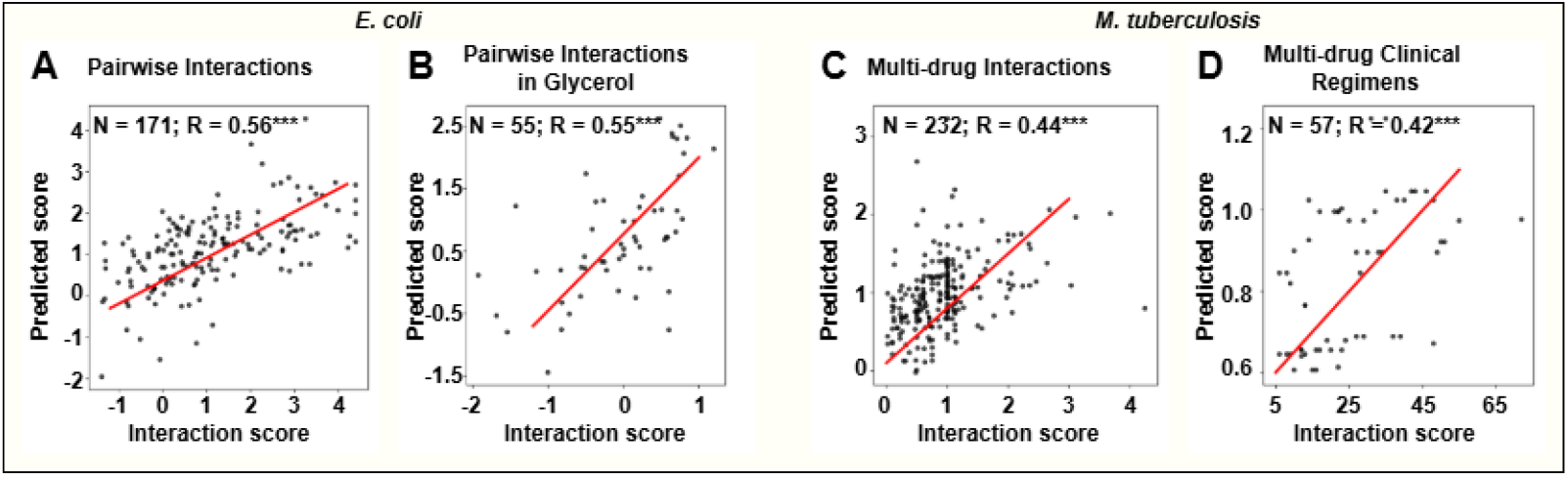
Scatterplot comparing predicted and actual interaction scores for (A) pairwise interactions, (B) pairwise interactions in glycerol for *E. coli*, (C) multi-drug interactions, and (D) multi-drug clinical regimens for *M. tuberculosis*.

**Supplementary Figure 3.**
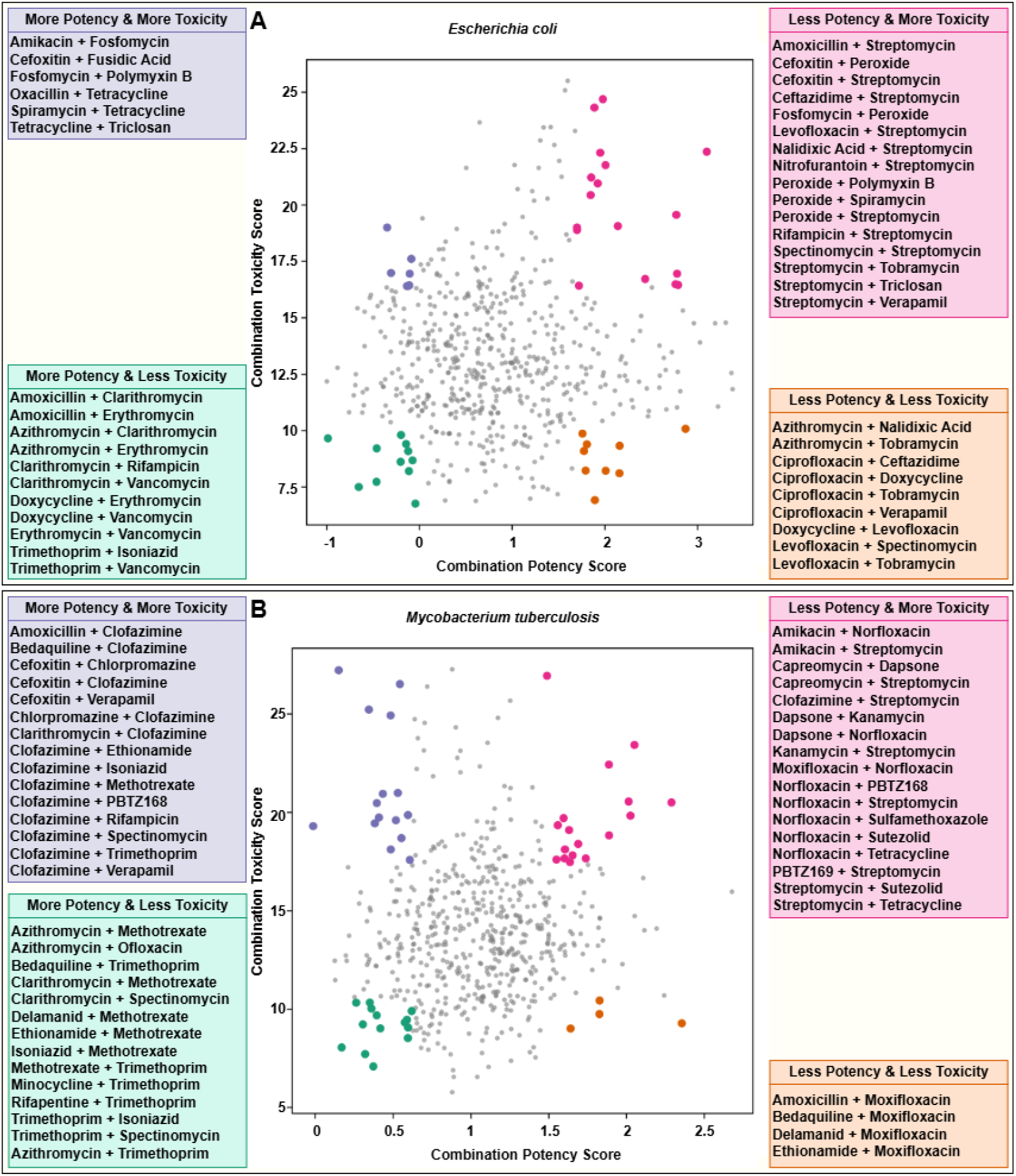
The drug combination landscape for **(A)** *E. coli* and **(B)** *M. tuberculosis*, highlighting all four categories of drug combinations based on lower and higher potency and toxicity, among 595 pairwise combinations from 35 drugs.

**Supplementary Figure 4.**
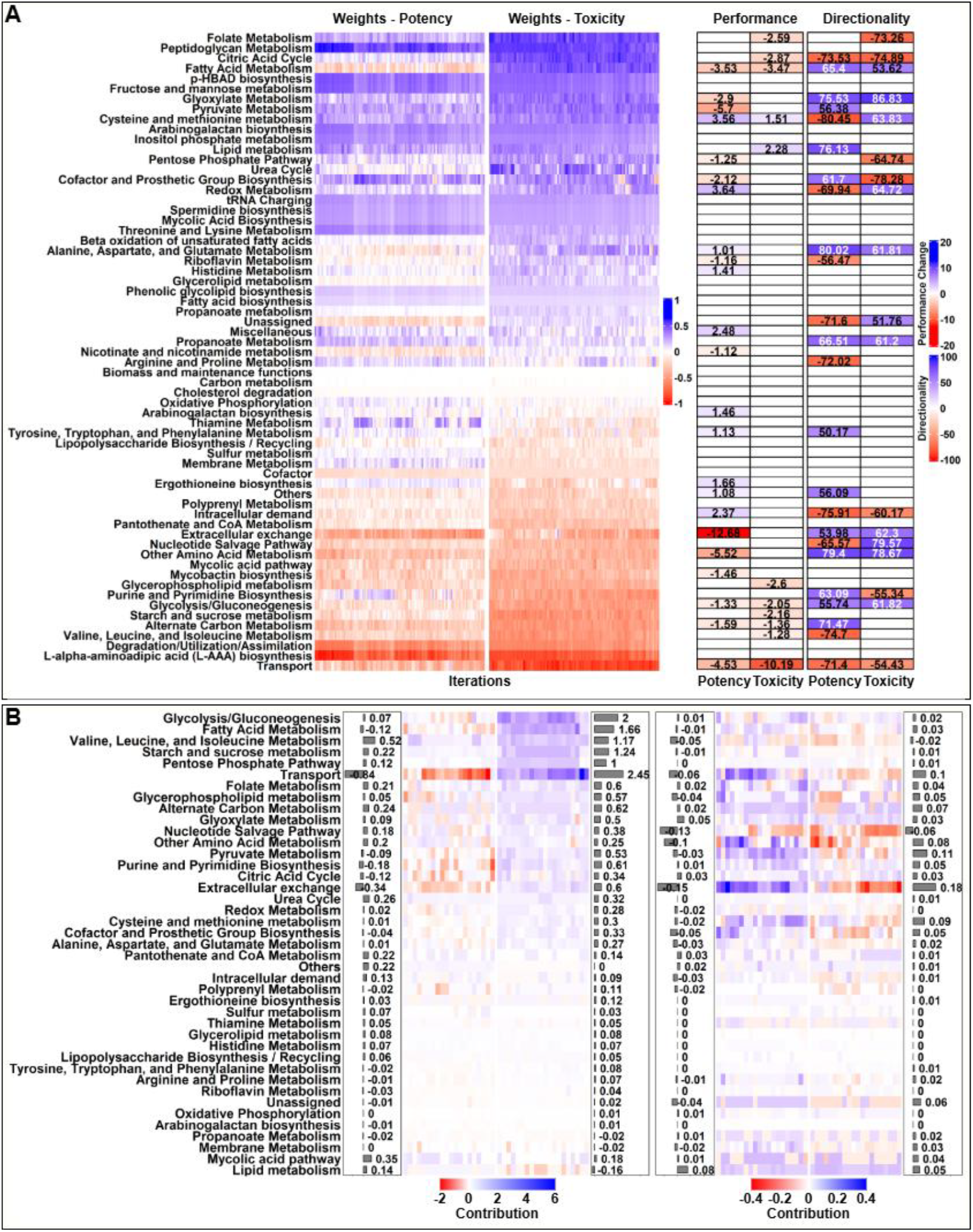
**(A)** Heatmap visualizes normalized weights for different pathways in *M. tuberculosis* models for drug combination potency and toxicity, showing the effect of removing pathway-specific features on model performance (note that changes within −1% to 1% are not considered significant and are omitted) and prediction directionality (note that changes within −50% to 50% are also omitted). **(B)** Cluster heatmap illustrating the weighted contributions of different pathways for the top and bottom 20 combinations regarding potency and toxicity for *M. tuberculosis*. The figure also includes bar charts displaying the mean contributions of individual pathways to the final prediction scores.

**Supplementary Figure 5.**
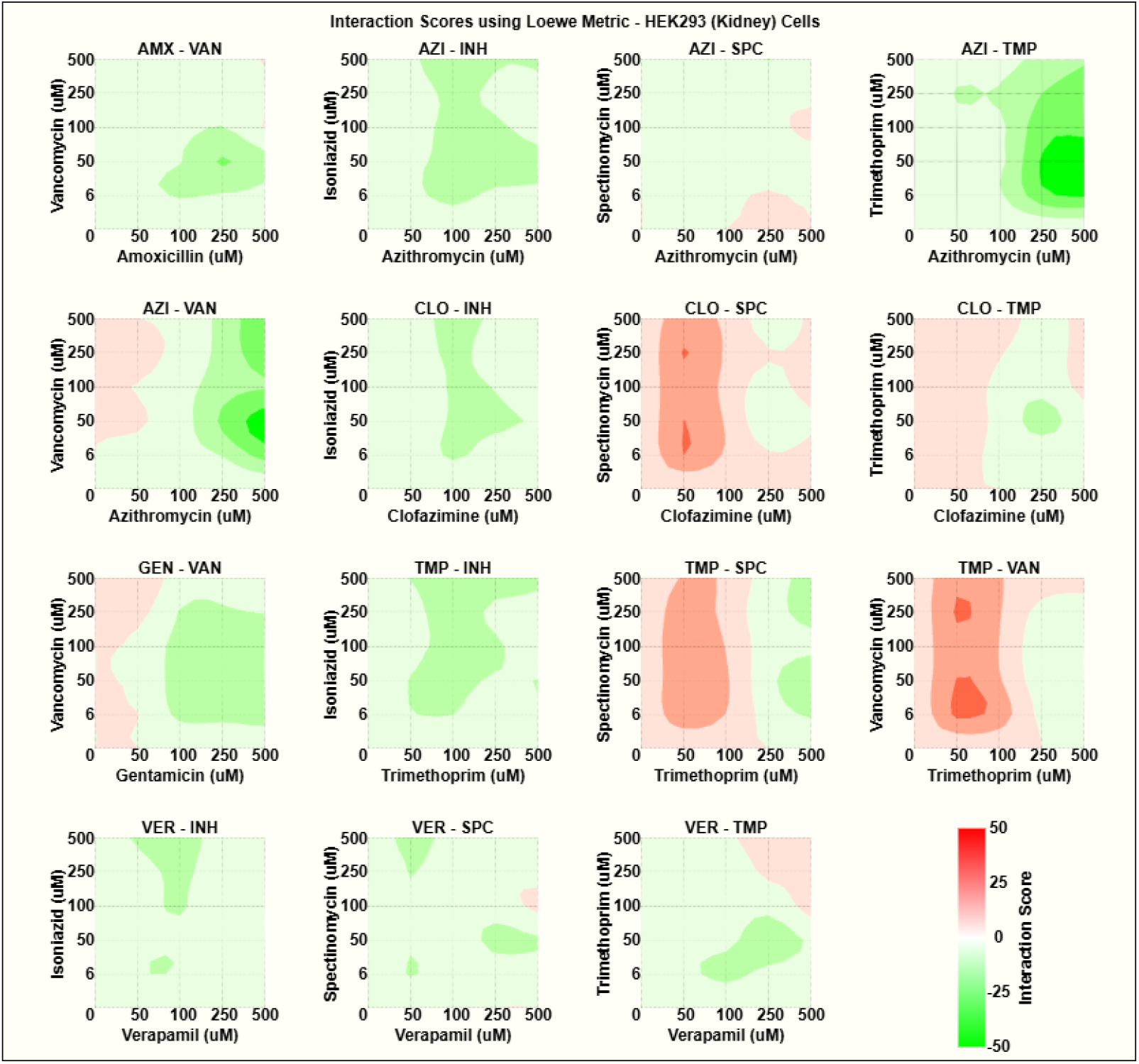
Heatmaps illustrating interaction (or synergy) scores at various dose levels for the selected 15 combinations using Loewe scoring metric in HEK293 cells.

**Supplementary Figure 6.**
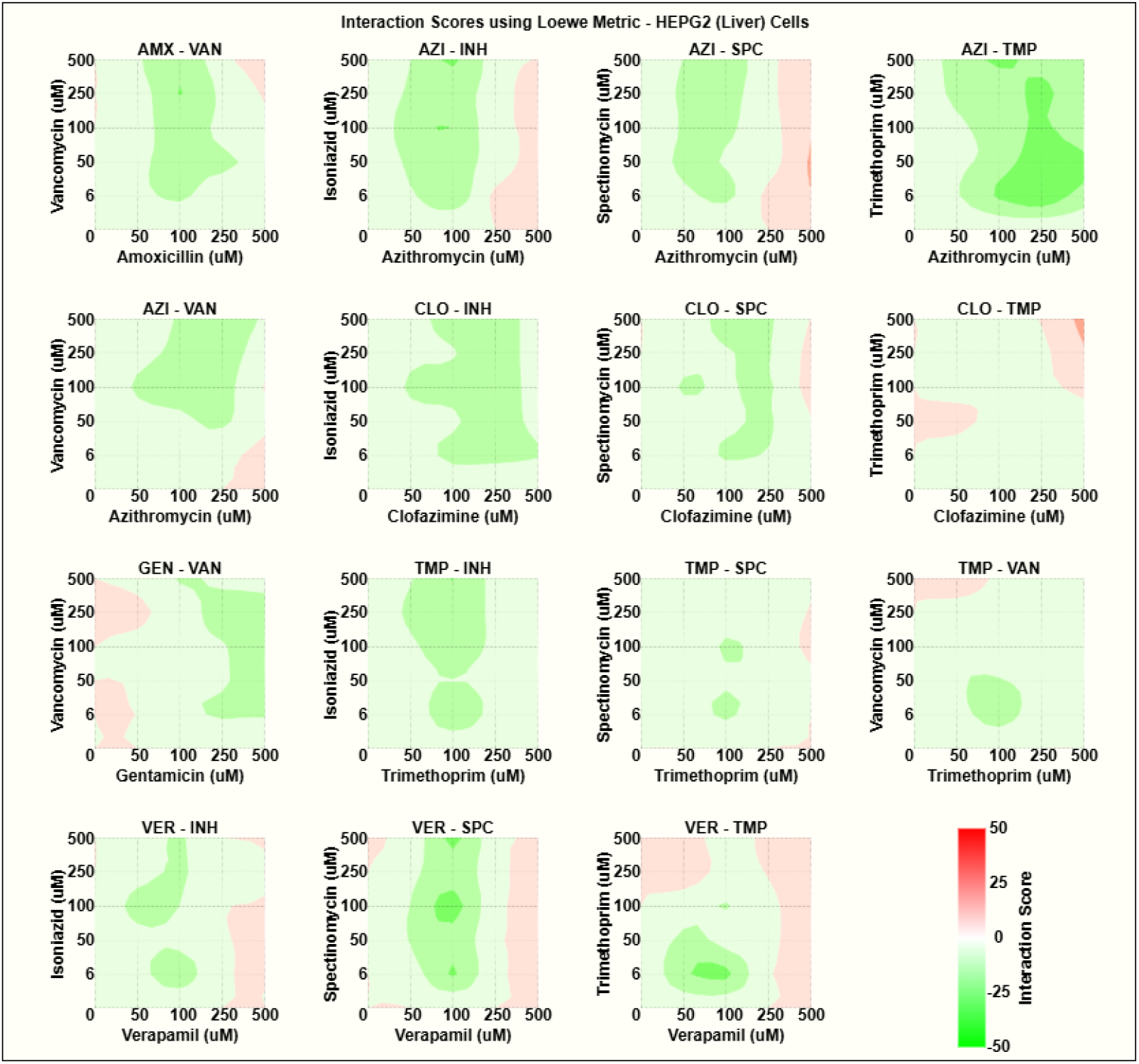
Heatmaps illustrating interaction (or synergy) scores at various dose levels for the selected 15 combinations using Loewe scoring metric in HEPG2 cells.

**Supplementary Figure 7.**
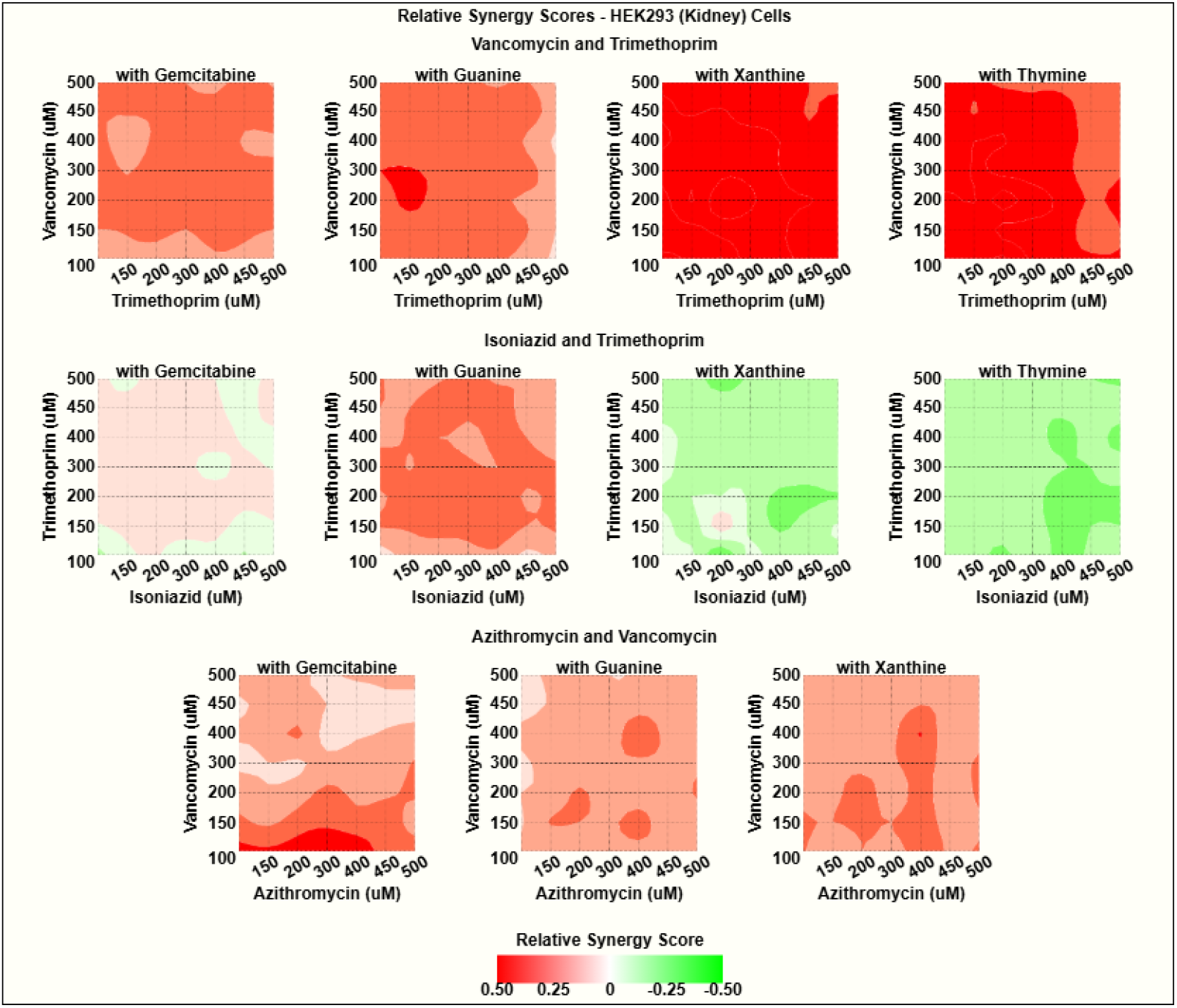
Heatmaps illustrating relative synergy scores of selected drug combinations in both the presence and absence of nucleotide metabolites in HEK293 cells, with positive scores (red) indicating decreased cell viability (or increased cell inhibition) when nucleotide metabolites are present.

**Supplementary Figure 8.**
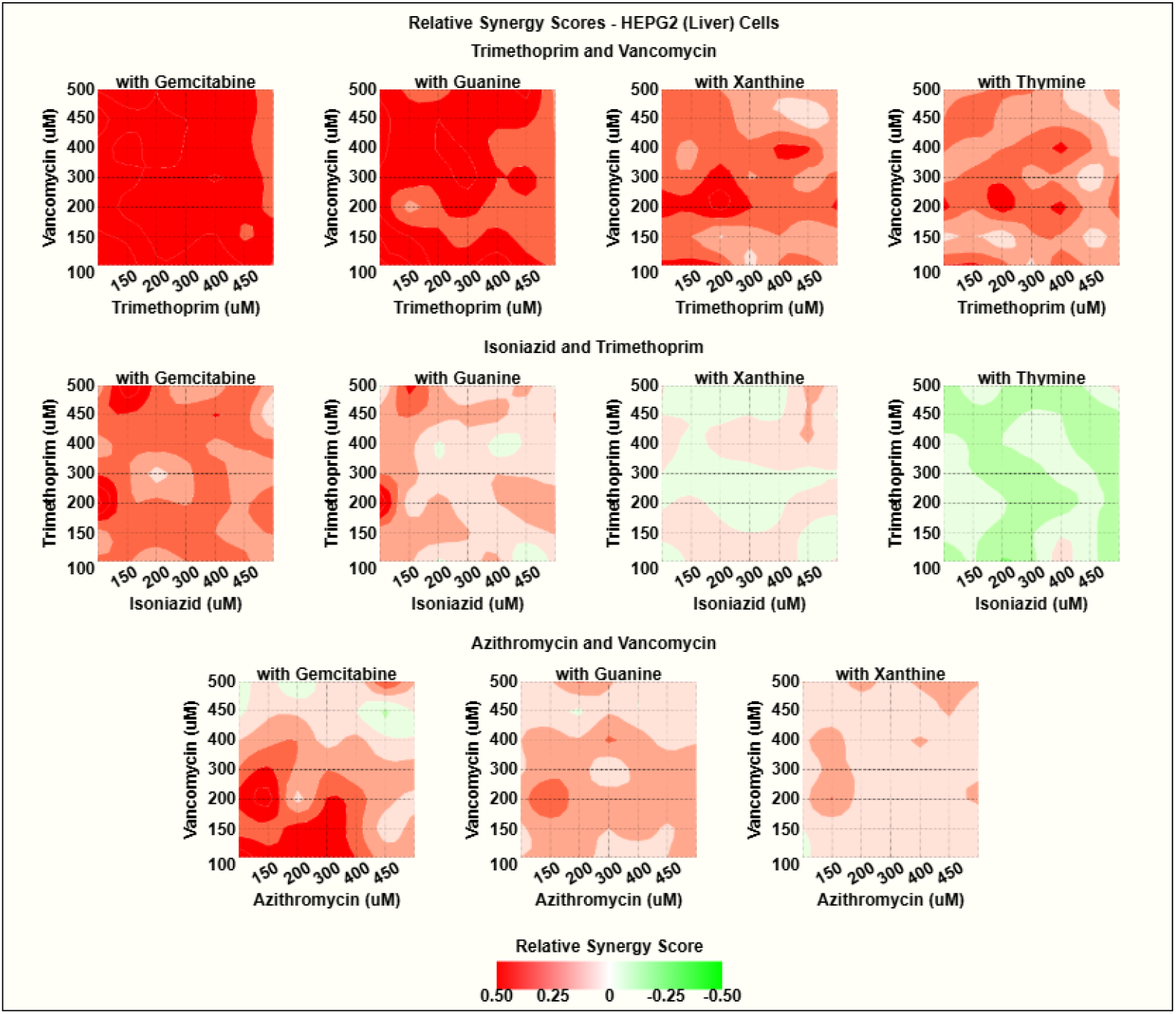
Heatmaps illustrating relative synergy scores of selected drug combinations in both the presence and absence of nucleotide metabolites in HEPG2 cells, with positive scores (red) indicating decreased cell viability (or increased cell inhibition) when nucleotide metabolites are present.

**Supplementary Figure 9.**
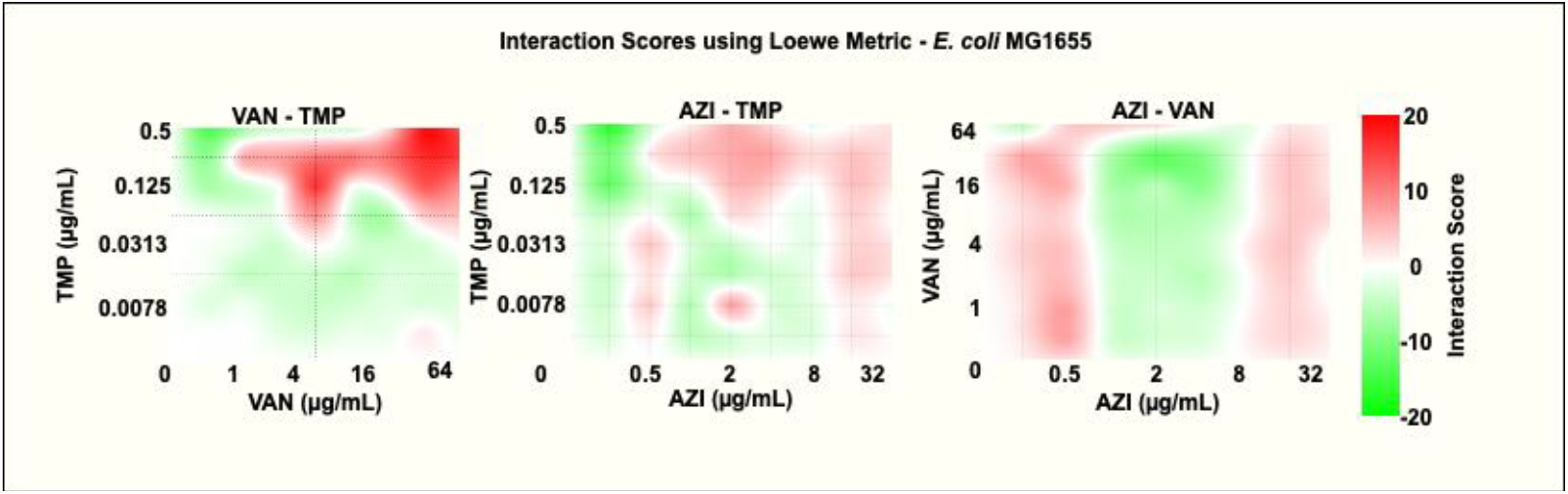
Heatmaps illustrating interaction (or synergy - indicated by the red color) scores at various dose levels for selected antibiotic combinations using Loewe scoring metric in *E. coli* MG1655.

**Supplementary Figure 10.**
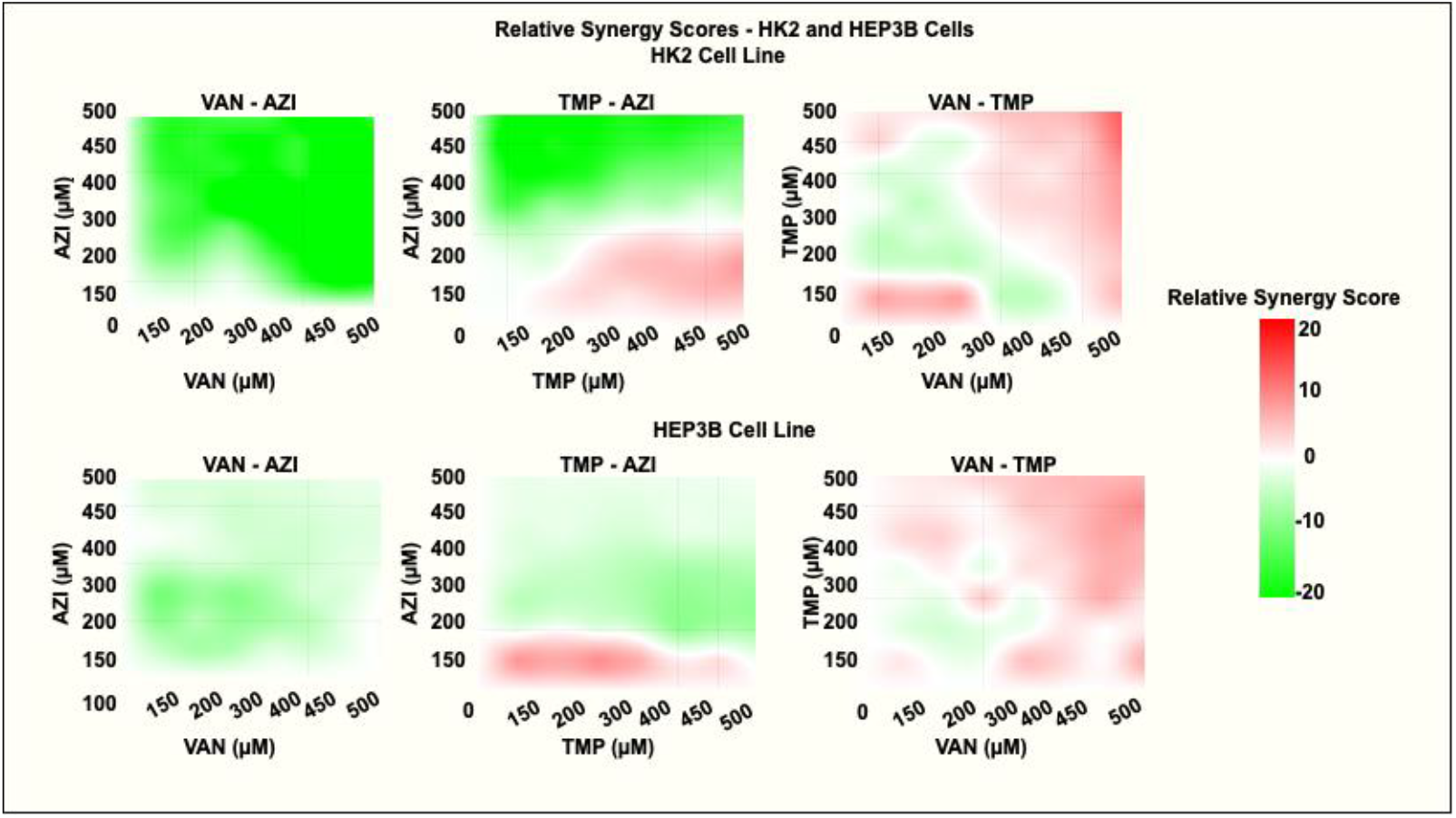
Heatmaps illustrating synergy scores (where synergy is indicated by the red color and antagonism indicated by the green color) at various dose levels for antibiotic combinations using the Loewe scoring metric. Combinations were evaluated in the HK2 and HEP3B cell lines.

